# Functional architecture of rare protein coding variation across 786,871 exomes

**DOI:** 10.64898/2026.10.16.26365771

**Authors:** Kangcheng Hou, Hui Li, Emily Nason, Hilary Finucane, Shamil Sunyaev, Luke J. O’Connor

**Affiliations:** Department of Biomedical Informatics, Harvard Medical School, Boston, MA; Program in Medical and Population Genetics, Broad Institute of MIT and Harvard, Cambridge, MA; Stanley Center for Psychiatric Research, Broad Institute of MIT and Harvard, Cambridge, MA; Analytic and Translational Genetics Unit, Massachusetts General Hospital, Boston, MA

## Abstract

Protein coding variants span the gamut of structural and functional effects on proteins, and they have similarly heterogeneous effects on traits. Their functional effects can be predicted and categorized, particularly using machine learning models, and integrating these predictors benefits gene discovery. However, it remains unclear how well known structural features and predicted functional scores explain the heterogeneous effects of protein coding variants on human complex traits. We introduce the Kernel Allelic Series Model (KASM), which integrates multiple functional annotations to form a kernel describing how allelic effects within a gene covary. We apply it to exome data from 26 traits and 786,871 individuals from UK Biobank and All of Us, integrating 35 annotations describing protein structure and predicting allelic effects. We estimate that rare missense heritability is at least 1.27±0.09% on average, 1.83±0.11% under our most flexible model, exceeding pLoF heritability (0.87±0.05%) and far exceeding missense burden heritability as previously estimated (0.54±0.04%). AlphaMissense is the single most enriched missense predictor, but a composite score doubles its enrichment. A series of structural features is enriched for heritability, from amino acid chemical features and structural motifs to residue contacts and protein-protein interaction interfaces. Examining pairs of variants, we find that most missense effects are modestly correlated, but an exception is when residues are close in 3D structure but distant in 1D sequence; such pairs also covary across species. Finally, we analyze individual genes and identify gene-specific enrichments, like a 95-fold enrichment among *PIEZO1* hydrogen bond residues, that nominate functional hypotheses.

## Introduction

Rare protein-coding variants are a major driver of gene discovery for common diseases and complex traits^1,2^, motivating the study of their functional effects. These effects are diverse, especially for missense variants, affecting protein stability^3^, protein-protein interactions^4^, post-translational modifications^5^, splicing^6^, and dosage^7^. Heterogeneity presents a statistical challenge: alleles with the most deleterious effects are extremely rare, and their rarity seemingly necessitates aggregation, yet effect-size heterogeneity dilutes aggregated burden statistics^8,9^. For this reason, the total contribution of missense variants to complex-trait heritability is unclear, and the interpretation of gene-aggregated missense associations is clouded by a lack of insight into the functional effects of individual alleles.

An opportunity exists to address this challenge by leveraging recent progress in machine learning and experimental approaches to protein structure and missense variant effect prediction. Such approaches both predict general deleteriousness^10–17^ and characterize specific molecular features: the position of an altered amino acid within a protein^18^, its effect on protein folding stability^19^, its effect on splicing^6^ and post-translational modifications^20^, and its interaction with other proteins and molecules^21,22^. Such annotations have been used to distinguish benign from damaging variants within known disease genes^23–25^ and to identify new gene-trait associations^26– 28^, but the opportunity extends much further. Within a gene, the pattern of functional and phenotypic effects across variants—its allelic series—can distinguish between hypotheses for the mechanism of an association^29,30^. Across the exome, the pattern defines protein coding functional architecture: what mechanisms are most prevalent; what aspects of protein biology are broadly relevant to traits; and against this background, what genes stand out individually. In common-variant association studies, a key approach has been to integrate association data with epigenomic and gene expression annotations, both at individual loci^31–33^ and across the genome^34,35^. For rare protein-coding variants, an analogous approach would integrate functional annotations describing how variants affect proteins, both exome-wide and within specific genes.

A functionally informed approach could also recover rare coding heritability missed by existing estimators. Burden Heritability Regression (BHR)^9^ estimates the component of rare coding heritability captured by prespecified burden statistics, but it does not account for within-gene heterogeneity. GREML-LDMS avoids gene-level aggregation, but its estimates become unreliable at minor allele frequencies below around 0.01%^36,37^. One challenge is that residual population stratification can bias heritability estimates^37^; another is that ultra-rare variants are carried by too few individuals for their contributions to be resolved without some amount of aggregation. A more flexible approach would allow variant effects which are neither identical nor independent, aggregating the effects of functionally similar alleles to the extent supported by the data.

Here, we introduce the Kernel Allelic Series Model (KASM), under which the effects of different alleles within the same gene covary according to their functional features. We integrate rare-variant association statistics from UK Biobank^38^ and All of Us^39^ with structural and functional annotations, including machine learning-derived predictions of variant effects and protein structure. We substantially revise previously published lower bounds on rare coding heritability^9^, identify enriched structural and functional features, quantify effect-size heterogeneity, and apply the same statistical approach to individual trait-associated genes.

## Results

### Methods overview

The premise of the Kernel Allelic Series Model (KASM) is that functionally similar alleles within the same gene, as predicted by relevant annotations, should have similar phenotypic effects. Mathematically, effect-size similarity is encoded by a kernel, or covariance matrix, which is calculated from the annotation data using a function fitted to the genetic data. Its diagonal elements describe the predicted magnitude of each effect; its off-diagonal elements describe how the effects of two variants covary. This covariance might be larger for similarly annotated alleles. For missense variants, these annotations could include deleteriousness scores^10,40,41^, allele frequency, and various features of amino acid substitutions, like whether they cause a change in polarity or disrupt a molecular interaction^18,23^ (**Methods**). The relevance of these annotations is learned from the data. This approach generalizes the burden-effect model, under which variant effects are perfectly correlated. A general kernel allows variant effects to be perfectly correlated, partially correlated, or independent, to the extent supported by the data.

Mathematically, KASM features an unobserved true effect-size covariance kernel, Σ, a model specifying some space of working kernels, *K*, and a distance metric which quantifies whether some kernel *K* ∈ *K* well approximates Σ (**Methods**). For the same Σ, different choices of *K* give rise to different distance-minimizing kernels *K*\* ∈ *K*. The estimand is *K*\*, not Σ, so the choice of model *K* affects both the estimator and the estimand. We consider a series of models incorporating different functional features, each defining different components of rare coding heritability (see analyses below).

KASM quantifies the *kernel heritability*, defined as the fraction of phenotypic variance explained by variant effects aligned with a specified kernel space (**Methods**). When the kernel is rank one, the kernel heritability reduces to the burden heritability. More generally, for an appropriate space of kernels, the kernel heritability is a lower bound on the total SNP-heritability (**Methods**). The *kernel heritability enrichment* of a subset of variants is defined as the fraction of heritability divided by the fraction of rare-variant heterozygosity (not the fraction of variants). This denominator accounts for allele-frequency differences; variants within an enriched annotation have higher per-allele effect sizes (**Methods**). We quantify this enrichment primarily among missense variants, separately within exome-wide significant gene-trait associations (“significant genes”) and other “subthreshold genes”.

KASM implements random-effects and fixed-effects estimators, suitable for analyzing the set of subthreshold genes and individual significant genes, respectively. Both operate on variant association statistics, within-gene linkage disequilibrium matrices, and functional annotations. The random-effects estimator (**Methods**) is a fast, summary-level implementation of Haseman-Elston regression^42,43^. It features diagonal kernels which absorb confounding due to residual population stratification, analogous to the BHR intercept^9^. It models the effects of subthreshold genes as draws from a shared distribution and excludes significant genes, so that the estimate is not driven by a few large effects. The fixed-effects estimator (**Methods**) treats each significant gene’s variant effects as fixed quantities without distributional assumptions, similar to ref.^44^. This estimator provides the resolution to interrogate individual genes and improves statistical power in exome-wide analyses. For exome-wide heritability estimation, we combine random-effects and fixed-effects estimates to obtain a mixed-effects estimate. The random- and fixed-effects estimators account for distinct sources of uncertainty across genes and sampled individuals (**Methods**).

### Simulations

We evaluated KASM in simulations with variant-level functional heterogeneity, gene-level differences in selective constraint, linkage disequilibrium (LD) computed from All of Us data, and excess variance in association statistics representing residual population stratification (**Methods**). Variant effects were simulated from a kernel derived from real functional annotations, with excess biological effect heterogeneity not explained by these annotations, and summary statistics were simulated conditional on LD. Fixing this data-generating model, we computed both true and estimated kernel heritabilities under four KASM models (none of which is exactly the data-generating model): the burden model of Weiner et al.^9^, under which variants are partitioned into discrete categories and aggregated (“BHR-like”); a more flexible model featuring several continuous-valued missense annotations (“KASM Baseline”; see below); a model further allowing distinct effects across exons within each gene (“KASM Baseline x Exon”); and a model which adds a heritability component for independent variant effects (“KASM Baseline + Indep.”). The true kernel heritability was smallest under the BHR-like model and largest under the KASM Baseline + Indep. model.

KASM accurately estimated the kernel heritability across simulation settings (**Figure 1a, Supplementary Figure 1** and **Supplementary Table 1**). The mixed-effects estimator reduced standard errors relative to treating all genes as random effects (**Supplementary Figure 2**). Ignoring LD within genes led to inflated heritability estimates (**Supplementary Figure 3**). KASM also accurately estimated heritability enrichment for enriched, depleted, and null missense variant subsets (**Figure 1b, Supplementary Figure 4**, and **Supplementary Table 2**). Enrichment estimates were well calibrated in both significant genes and subthreshold genes, supporting the use of both estimators. In the same simulations, KASM also accurately estimated effect concordance across within-gene variant pairs for both missense and pLoF variants (**Supplementary Figure 5**).

**Figure 1.**
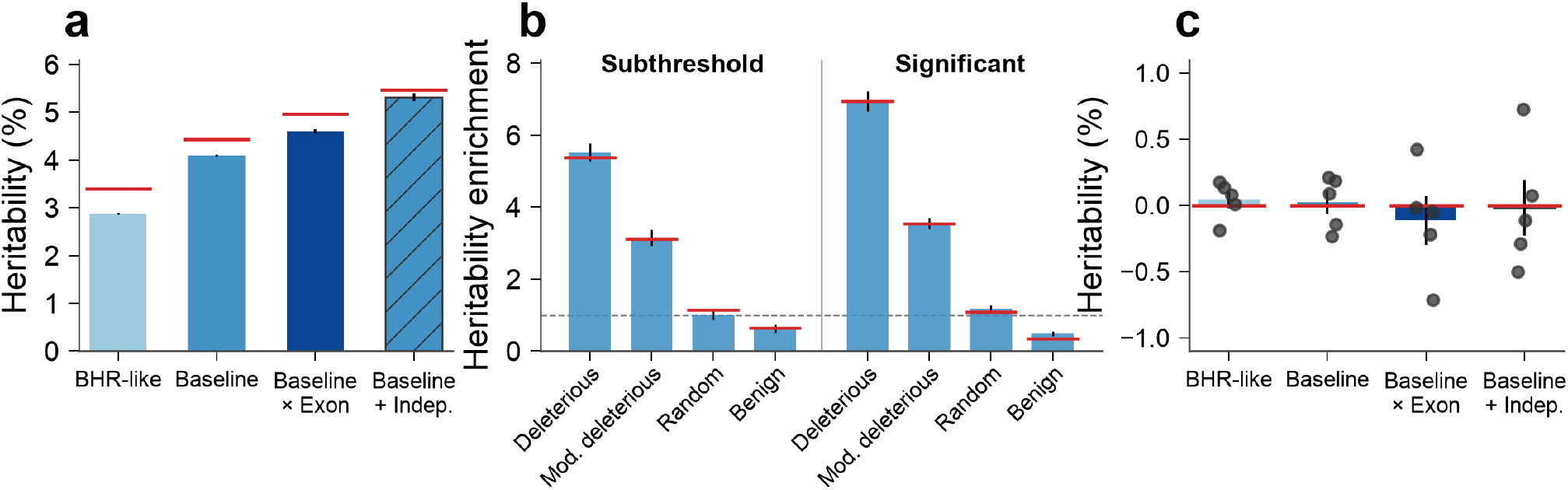
Simulations. **(a)** Heritability estimates under the BHR-like, KASM Baseline, KASM Baseline x Exon and KASM Baseline + Indep. models. Bars show means across 100 replicates, and red segments show the corresponding simulated estimands. **(b)** KASM Baseline heritability enrichment of missense annotation subsets in subthreshold and significant genes. Each set contains 2% of missense variants within each gene, ranked by AlphaMissense: deleterious (0-2%), moderately deleterious (9-11%), random and benign (98-100%). Bars show estimates across 100 replicates, and red segments show simulated enrichment. **(c)** Heritability estimates in null simulations with common-variant effects but no rare coding heritability, meta-analyzed across All of Us EUR-, AFR- and AMR-like cohorts. Bars show means and points show individual estimates across five replicates. Error bars show standard errors.

We additionally evaluated KASM using phenotypes simulated in the All of Us “*All by All*” data release with zero kernel heritability but nonzero common-variant heritability^39^. Estimates were near-zero, as expected, across models and ancestry groups (**Figure 1c** and **Supplementary Table 3**). These individual-level simulations show that KASM avoids genetic confounding due to population structure and LD.

### Functionally informed kernel heritability

We applied KASM to exome association statistics for 16 UK Biobank traits and 19 All of Us traits (26 distinct, well-powered continuous traits), including up to 394,841 UK Biobank and 392,030 All of Us individuals (**Supplementary Table 4**). We analyzed approximately 17K genes and 15 million rare coding variants (MAF < 0.1%; including singletons), classified as pLoF, missense, or synonymous variants. For missense variants, we compiled 15 quantitative deleteriousness scores, including PolyPhen-2^10^, AlphaMissense^14^, and ESM-1v^12^, which were only partially correlated (**Figure 2a** and **Supplementary Table 5**); and 20 structural and functional annotations spanning primary sequence, secondary and tertiary structure, and protein-protein interaction interfaces (**Supplementary Table 6**). We meta-analyzed kernel heritability estimates across continuous traits. We separately analyzed four UK Biobank binary disease traits and did not include them in subsequent analyses because of their low observed-scale heritability (**Methods** and **Supplementary Table 7**).

**Figure 2.**
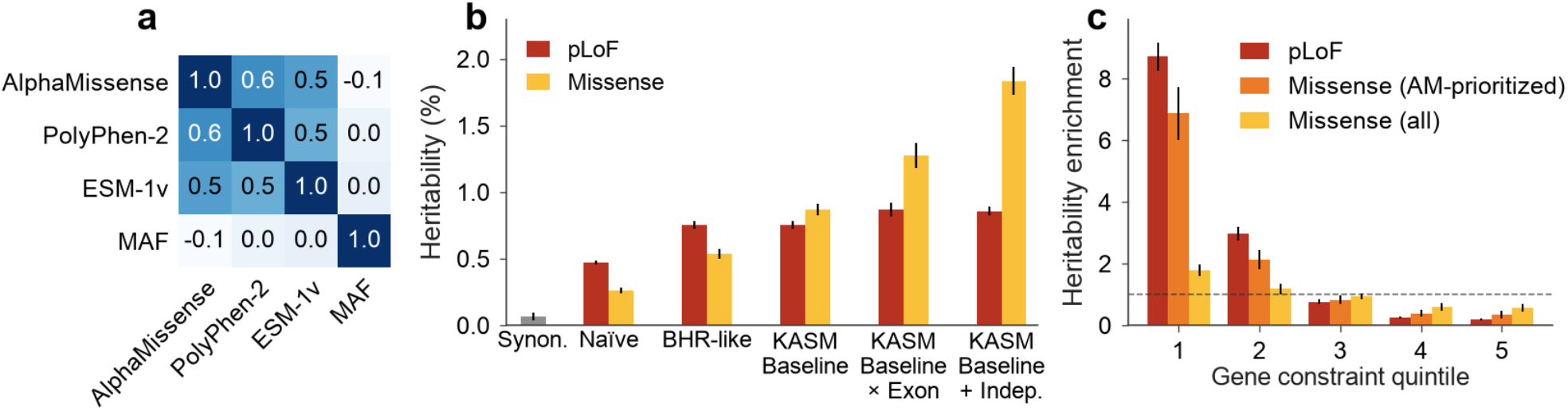
Functionally informed estimation of rare coding heritability. **(a)** Spearman correlations among the KASM Baseline missense annotations and minor allele frequency (MAF).**(b)**Rare coding heritability of pLoF and missense variants under the Naïve (constant burden), BHR-like (binarized PolyPhen-2 stratified by MAF), KASM Baseline (PolyPhen-2, ESM-1v and AlphaMissense stratified by MAF), KASM Baseline x Exon and KASM Baseline + Indep. models, and of synonymous variants (Synon.). **(c)** Heritability enrichment across gnomAD pLoF observed/expected quintiles, from most (1) to least (5) constrained, for pLoF, AlphaMissense-prioritized (AM-prioritized) missense and all missense variants. Estimates are meta-analyzed across UK Biobank and All of Us traits. Error bars show standard errors.

We estimated kernel heritability under several models, producing a sequence of lower-bound estimates for rare missense heritability (**Figure 2b** and **Supplementary Table 7**). Synonymous heritability was near zero (0.07±0.03%). Under a “naïve” burden model, which assigns one burden effect to all missense variants in each gene, the missense kernel or burden heritability was 0.26±0.02% (under this model, kernel and burden heritability are equivalent). Under a “BHR-like” model, which partitions missense variants by MAF and a binarized PolyPhen-2 score, the kernel or burden heritability was 0.54±0.04%, still smaller than the pLoF burden heritability (0.75±0.03%). Under a model containing three quantitative deleteriousness scores (AlphaMissense, ESM-1v, PolyPhen-2) stratified by MAF, termed “KASM Baseline”, the missense kernel heritability was 0.87±0.05%, slightly surpassing pLoF heritability. Stratification by MAF was important, as ultra-rare variants had larger per-allele effects than higher-frequency variants with similar predicted deleteriousness (**Supplementary Figure 6**). Adding additional missense annotations had little effect (**Supplementary Figure 7**), likely because these annotations were redundant or contained very few variants.

Next, to capture additional effect-size heterogeneity that is poorly predicted by deleteriousness scores, we fitted a “KASM Baseline x Exon” model, which allows effects to vary across exons, and “KASM Baseline + Indep.”, which allows variants to have independent effect-size residuals, particularly at higher allele frequencies (**Figure 2b, Supplementary Table 7**). Missense kernel heritability increased from 0.87±0.05% under KASM Baseline to 1.27±0.09% under KASM Baseline x Exon and 1.83±0.11% under KASM Baseline + Indep. The increase under the exon model may reflect splicing-related effects or any other source of distance-dependent heterogeneity. The corresponding pLoF estimates were 0.75±0.03%, 0.87±0.05%, and 0.86±0.03%. Among these models, KASM Baseline yielded the strongest statistical evidence for nonzero kernel heritability (*P*=7×10^−82^ and 2×10^−152^ for missense and pLoF variants), suggesting a useful balance between model flexibility and estimation precision, and is used as a default baseline model in subsequent sections (**Supplementary Table 7**).

The KASM Baseline + Indep. model, which yields the highest kernel heritability, relies on additional assumptions in order to separate biological effects from confounding. Like the other models, it includes “overdispersion” kernels intended to absorb confounding effects, and these confounding effects are assumed to be uncorrelated (**Methods**). Unlike other models, it also features a biological kernel featuring uncorrelated effects, raising the question of identifiability. To distinguish confounding effects from independent biological effects, we assume that the former are equally strong for missense and synonymous variants at the same allele frequency (**Supplementary Figure 8**), and moreover that they do not vary with gene-level selective constraint. These assumptions are plausible but not fully testable. An additional limitation of this analysis is that the independent-effects kernel is not frequency stratified; for this reason, the kernel heritability under this model remains a lower bound on the total missense heritability.

The additional missense heritability explained by the expanded models was concordant in both significant and subthreshold genes (**Supplementary Figure 9**); for the KASM Baseline + Indep. model, this concordance is reassuring because the ratio of biological vs. confounding effects should be much higher within significant genes. Some specific gene-trait associations exhibited differences across models, including a pLoF association between *TET2* and platelet count, for which heterogeneous pLoF effects probably reflect somatic clonal expansion^45,46^ (**Supplementary Figure 10**). As a negative control, synonymous heritability remained near zero under these models (**Supplementary Table 7**).

Previous burden heritability estimates^9^ were ~2x higher for pLoF variants than for missense variants. The primary difference in our analysis is that a more flexible model captures more missense heritability; when fitted to the same set of 16 UK Biobank traits, KASM “BHR-like” and BHR produced concordant missense burden heritability estimates (**Supplementary Figure 11**). An additional difference is that for pLoF variants, KASM produced lower estimates than BHR, by about 11% (**Supplementary Figure 11**). This difference is related to LD among pLoF indels, which are sometimes represented by multiple correlated variant records. KASM accounts for this using within-gene LD matrices, whereas the previous analysis^9^ used a correction factor assuming that variants in different functional categories have similar amounts of LD. We verified that, without LD correction, KASM and BHR produced concordant pLoF heritability estimates (**Supplementary Figure 11**).

We examined how rare coding effects vary with gene-level selective constraint estimated from pLoF variants in gnomAD^47^ (**Figure 2c** and **Supplementary Table 8**). pLoF variant effects were 8.7-fold enriched in the most constrained gene quintile. Missense variants in aggregate were only 1.8-fold enriched, apparently suggesting that pLoF variant constraint is weakly relevant to missense variant effects; however, AlphaMissense-prioritized missense variants were 6.9-fold enriched, approaching the pLoF enrichment (**Methods**). Without conditioning on deleteriousness, effect-size heterogeneity among variants combines with the effect of negative selection to dilute, or flatten^48^, the heritability enrichment of constrained genes: deleterious variants are depleted, and segregating variants are biased toward milder effects, reducing apparent differences between genes (**Supplementary Figure 12**). A related phenomenon is that enrichment in constrained genes is greatly attenuated when measured in units of heritability per gene (we instead measure effect-size enrichment); Spence et al.^49^ reported that constrained genes are unenriched for pLoF heritability per gene, but the discordance with our results is superficial.

We performed two secondary analyses. First, we assessed potential bias from residual population stratification aligned with functional annotations (**Methods**). We determined that their contribution was minimal for most traits (below 0.1% of phenotypic variance; **Supplementary Figure 13**), concordant with Weiner et al.^9^. Second, we added the genotype covariance matrix inverse as an overdispersion kernel, motivated by the theoretical possibility that the kernel heritability could exceed the total heritability (under conditions described in **Methods**). This addition had little effect, supporting the interpretation of the kernel heritability as a lower bound on the total heritability (**Supplementary Figure 14**).

### Benchmarking missense deleteriousness predictors

We next evaluated how well individual missense deleteriousness predictors prioritize trait-associated rare missense variants. We compared 15 predictors, including the three used in the KASM Baseline model (PolyPhen-2, ESM-1v, and AlphaMissense). For each predictor, we ranked missense variants within each gene and estimated heritability enrichment for variants in the top decile. This approach measures a predictor’s ability to distinguish variants with larger versus smaller phenotypic effects in the same gene, rather than prioritizing variants in genes with larger overall phenotypic effects (**Methods**).

Top-decile variants from every deleteriousness predictor were enriched for missense heritability in both significant and subthreshold genes (**Figure 3** and **Supplementary Table 9**). Predictors based on neural-network representations of protein sequence and structure, including AlphaMissense, MisFit-S and PrimateAI-3D, showed the strongest enrichment. AlphaMissense and MisFit-S additionally leverage human population variation through allele-frequency information for supervised training and allele-count-based estimates of selection, respectively. The stronger performance of neural-network-based predictors relative to other traditional and ensemble predictors is consistent with previous benchmarks using functional assays and clinical variant classifications^13,14^.

**Figure 3.**
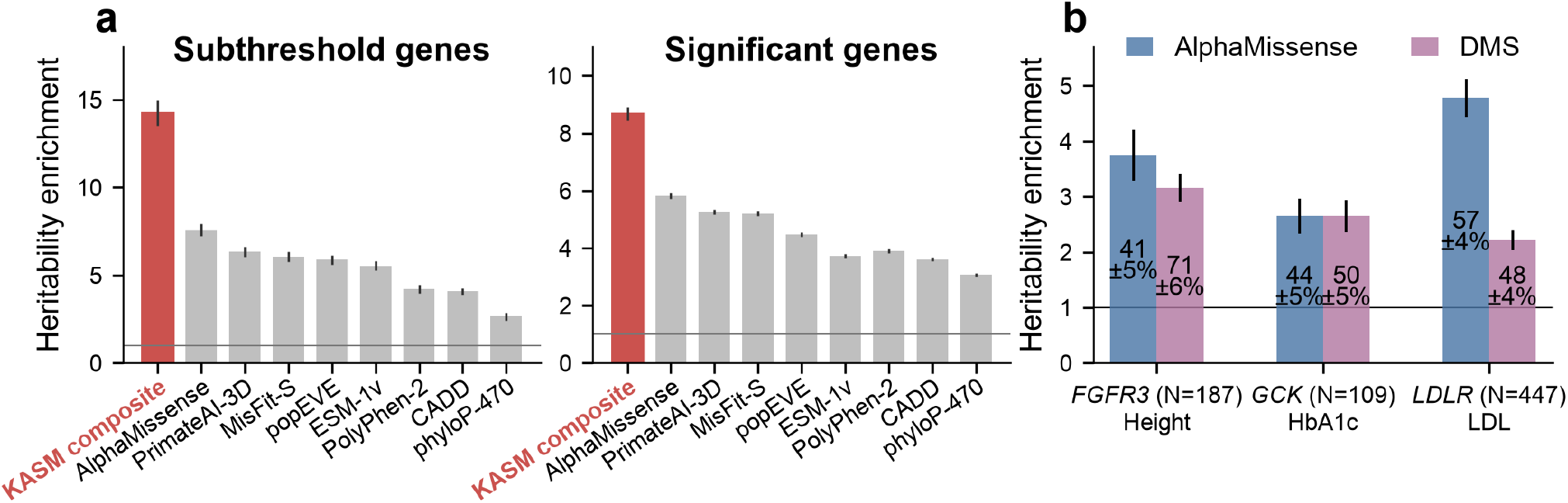
Benchmarking missense variant prioritization. **(a)** Heritability enrichment of the top decile of missense variants, ranked within genes by the KASM composite score or by selected deleteriousness predictors, in subthreshold and significant genes, meta-analyzed across UK Biobank and All of Us traits; all predictors are reported in **Supplementary Table 9. (b)** Top-quartile heritability enrichment for AlphaMissense and deep mutational scanning (DMS) in three significant gene-trait associations in UK Biobank. N denotes the number of variants with benchmark scores, and labels give the percentage of heritability among these variants (± standard error) in the top quartile. Error bars show standard errors.

We then constructed a KASM composite score from MAF-stratified deleteriousness predictors, with weights estimated using an even-odd chromosome split to avoid overfitting (**Methods**). The composite score outperformed every individual predictor (**Figure 3** and **Supplementary Table 9**): its top-decile variants were 14.2±0.7-fold enriched in subthreshold genes and 8.7±0.2-fold enriched in significant genes, compared with 7.6-fold and 5.8-fold for AlphaMissense. Much of the improvement came from MAF stratification; the top composite decile prioritized variants with larger per-allele effect magnitudes and was therefore strongly depleted of higher-frequency rare variants (**Supplementary Figure 15**). With a different definition of enrichment, prioritizing heritability per variant rather than effect size magnitude, an optimal composite score prioritizes higher-frequency variants instead (**Supplementary Table 9** and **Methods**). The lower enrichment in significant genes partly reflected genes whose trait-associated variants were poorly prioritized by deleteriousness predictors. One example is the *GPT*-alanine aminotransferase (ALT) association (2.5±0.2-fold for AlphaMissense). *GPT* encodes the cytosolic ALT1; variants reducing GPT activity can strongly alter measured ALT without large fitness effects, which may make them difficult for general deleteriousness predictors to prioritize.

These results indicate that, among missense variants observed in population-scale exome data, trait-relevant deleterious missense variants are best prioritized by combining functional predictors with allele frequency information, consistent with improved power for gene discovery^26,28^. Allele frequencies are highly relevant when analyzing inherited variation, for which observed alleles are sampled in proportion to their population frequencies; by contrast, most de novo or experimentally generated mutations are absent from population reference datasets (**Discussion**).

We compared computational predictors with deep mutational scanning (DMS) assays, which experimentally measure molecular variant effects in specific contexts^50^. Four significant gene-trait associations had sufficient overlap between DMS measurements and observed exome variants. Within the common set of variants for each assay, we compared heritability enrichment for top-quartile variants prioritized by DMS or by computational predictors (**Methods**).

The relative performance of DMS and computational predictors varied across gene-trait associations (**Figure 3, Supplementary Figure 16** and **Supplementary Table 10**). For *FGFR3* and height, top-quartile variants from a kinase-domain activation assay^51^ had enrichment comparable to AlphaMissense (3.2±0.3-fold vs. 3.7±0.5-fold), while accounting for more heritability because they had higher allele frequencies (71±6% vs. 41±5%). For *GCK* and HbA1c, a glucokinase activity assay^52^ performed similarly to AlphaMissense, whereas AlphaMissense outperformed an LDL-uptake assay^53^ for *LDLR* and LDL cholesterol. Results were similar for *MC4R* and BMI using an MC4R functional assay^54^. DMS assays therefore captured a component of effect-size heterogeneity within trait-associated allelic series, but apparently not to a greater degree than AlphaMissense, at least across four gene-trait pairs. Computational predictors have the advantage that they can be applied to any variant in any gene at little incremental cost, while experimental assays have the advantage that their specific readout can be mechanistically interpretable (**Discussion**).

### Structural and functional enrichments

We next asked where within proteins rare missense heritability is concentrated. Proteins are organized hierarchically, from the primary amino acid sequence to secondary structural motifs, tertiary folding within a single chain, and the formation of protein complexes via protein-protein interactions. We examined 20 structural and functional annotations spanning this hierarchy. Enrichment estimates were concordant between exome-wide significant and subthreshold genes, with smaller standard errors in significant genes; we report estimates from significant genes below (**Figure 4** and **Supplementary Table 11**).

**Figure 4.**
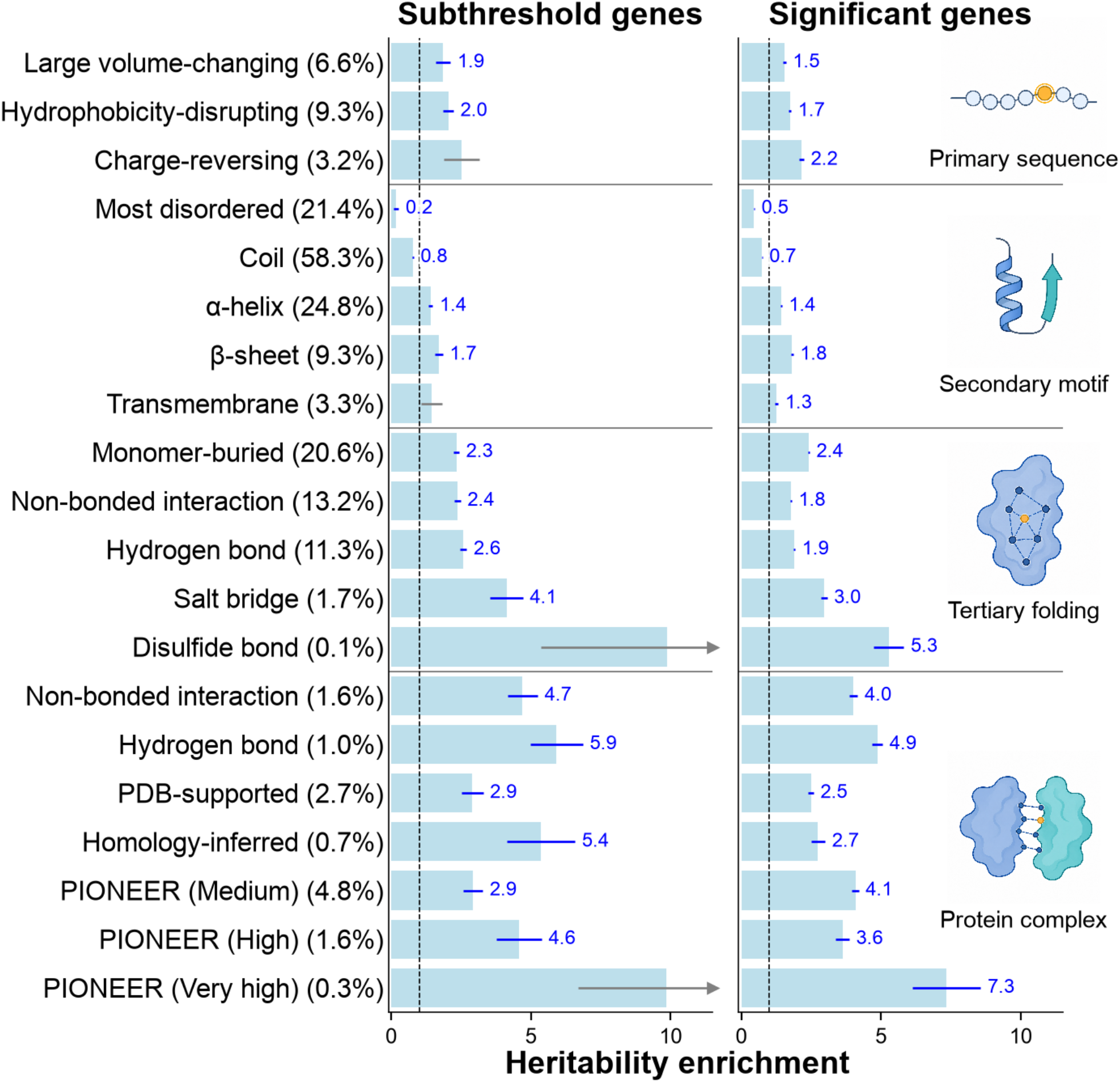
Missense heritability enrichment across levels of protein organization. Heritability enrichment for 20 structural and functional missense annotations in subthreshold and significant genes. Tertiary-folding contacts are within a protein chain; protein-complex contacts and interfaces are between chains. Parentheses give each annotation’s proportion of rare missense heterozygosity across all genes. Blue error bars and labels (enrichment estimates) denote annotations passing two-sided Bonferroni correction for 20 annotations; arrowheads denote values beyond the plotted range. Estimates are meta-analyzed across UK Biobank and All of Us traits. Error bars show standard errors.

At the level of the primary sequence, substitutions between amino acids with divergent physicochemical properties were enriched for missense heritability. Charge-reversing substitutions (negative-to-positive or positive-to-negative) showed 2.2±0.09-fold enrichment (explaining 9% of heritability vs. 4% of heterozygosity). Hydrophobicity-disrupting substitutions (hydrophobic-to-polar or polar-to-hydrophobic) showed 1.7±0.04-fold enrichment (20% vs. 11%), and large amino-acid volume changes (absolute volume change > 90Å^3^) showed 1.5±0.06-fold enrichment (10% vs. 7%).

At the level of secondary structure, variants in regular structural elements were enriched, whereas variants in disordered regions were depleted. Variants in α-helices and β-sheets were 1.4±0.02-fold and 1.8±0.04-fold enriched, respectively. In contrast, variants in highly disordered regions were depleted, with a 0.5±0.02-fold enrichment, consistent with stronger effects in structurally rigid regions.

At the level of tertiary structure, buried residues and intrachain contacts were enriched, consistent with their potential to disrupt protein stability. Variants in buried residues (relative solvent accessibility < 0.25) showed 2.4±0.02-fold enrichment (59% of heritability from 24% of heterozygosity). Intrachain contacts were also enriched, including non-bonded contacts at 1.8±0.02-fold, hydrogen-bond residues at 1.9±0.03-fold, and salt bridges at 3.0±0.1-fold. Disulfide-bond residues showed particularly strong enrichment (5.3±0.5-fold), although they represented only 0.28% of heterozygosity.

The strongest enrichments occurred at protein-protein interaction interfaces. Residues forming interchain hydrogen bonds were enriched 4.9±0.2-fold. Interfaces supported by Protein Data Bank structures were enriched 2.5±0.09-fold, homology-inferred interfaces 2.7±0.3-fold, and the highest-confidence PIONEER-predicted interfaces^21^ 7.3±1.2-fold, although this category represented only 0.08% of heterozygosity.

Broadly, these results demonstrate that well-known structural features, many of which are also enriched for Mendelian disease mutations^21,23,25^, are generically relevant within trait-associated allelic series. This suggests that trait-associated missense alleles often have unexotic structural effects on the stability of proteins and especially protein complexes. When investigating missense associations within a gene, a logical starting point is to overlay structural features; whatever change in protein structure is phenotypically relevant, it likely aligns or at least correlates with some feature that is already well annotated.

### Context-dependent variant effects

We next tested whether certain missense annotations interact to amplify heritability enrichments. Specifically, we asked whether variants with higher-level structural features (e.g., hydrogen-bond residues) had markedly different enrichments depending on the particular amino acid substitution (e.g., whether the substitution was charge-reversing) (**Methods**). We analyzed the three primary-sequence substitution annotations from **Figure 4** together with the 17 other, structural annotations and identified 29 annotation pairs for which the heritability enrichment of the structural annotation differed significantly depending on the chemical effect of the amino acid substitution (**Supplementary Figure 17** and **Supplementary Table 12**).

The strongest interaction involved charge-reversing substitutions at interchain hydrogen-bond residues, which had 27±2-fold heritability enrichment, compared with 3.9±0.2-fold for other substitutions at these residues (**Figure 5a**). Neither hydrophobicity-disrupting nor large-volume-changing substitutions showed a comparable interaction (**Supplementary Figure 17**). Charge-reversing substitutions were also strongly enriched at intrachain hydrogen bonds (6.9±0.4-fold vs. 1.8±0.03-fold) and intrachain salt bridges (9.3±0.8-fold vs. 2.6±0.1-fold). At residues buried within protein monomers, all three primary-sequence substitution classes were enriched (**Supplementary Figure 17**), suggesting that buried residues can perturb protein structure in a variety of ways; for example, volume-changing substitutions could cause cavity formation or overpacking (**Figure 5b**). Charge-reversing substitutions at buried H-bond residues were 13±1-fold enriched, compared with 2.7±0.05-fold for other substitutions (**Figure 5c**); such variants might be especially destabilizing due to an inability to form compensatory hydrogen bonds with water. Broadly, these estimates imply that missense effects depend upon the interaction between amino acid-level biochemical properties and higher-level structural features.

**Figure 5.**
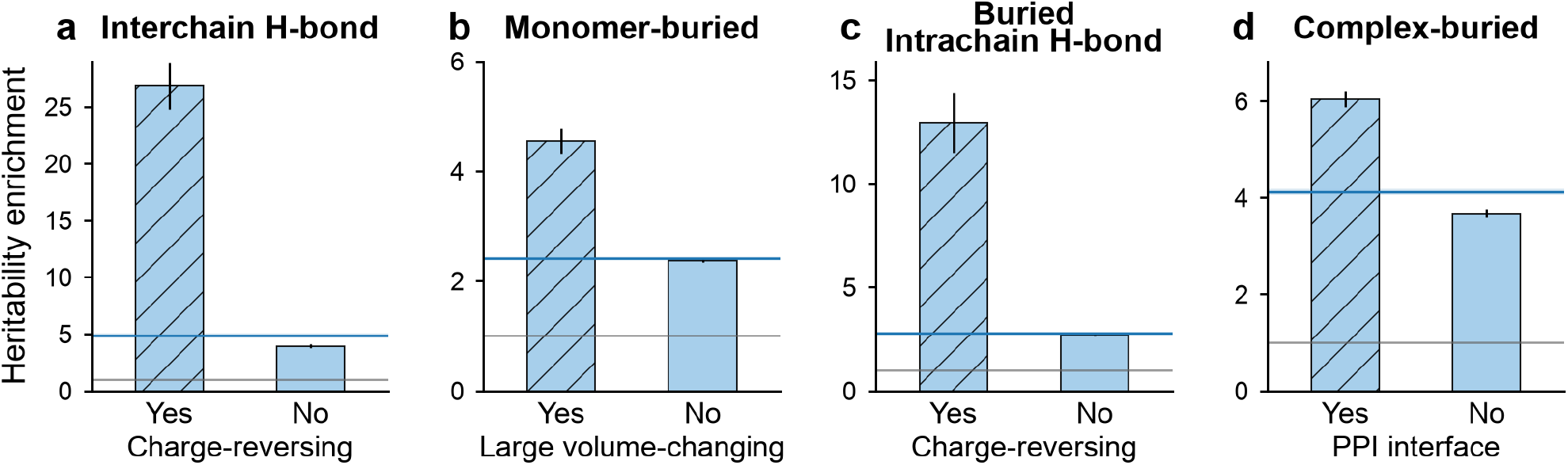
Missense heritability enrichment by amino acid substitution class. Heritability enrichment of missense variants in significant genes, within (Yes) and outside (No) a substitution class in a structural context. **(a)** Charge-reversing substitutions at interchain hydrogen-bond residues. **(b)** Large volume-changing substitutions at monomer-buried residues. **(c)** Charge-reversing substitutions at monomer-buried intrachain hydrogen-bond residues. **(d)** Protein-protein interaction (PPI) interface residues among complex-buried residues. Blue lines and bands show enrichment ± standard error for the full context. Estimates are meta-analyzed across UK Biobank and All of Us traits. Error bars show standard errors.

Missense variants can disrupt protein complexes either indirectly, by destabilizing the folded structure of a constituent protein, or directly, by altering a protein-protein interaction interface^4,55^. Both mechanisms are common among Mendelian disease mutations, which are enriched in buried residues and at protein-protein interfaces^4,25^. We annotated variants by whether their residues were buried in a protein complex and then compared interface and non-interface residues within this complex-buried variant set. Within complex-buried variants, interface residues had significantly higher enrichment than non-interface residues (6.0±0.2-fold vs. 3.7±0.1-fold; **Figure 5d**). Thus, the enrichment we observed at protein interfaces is not explained by the generic enrichment of buried residues.

### Effect concordance of variant pairs

Missense variants at nearby residues may perturb the same structural or functional element and therefore produce more concordant trait effects than distant variants. For example, pathogenic missense variants are known to cluster in three dimensional structures of some proteins^56–58^, but whether spatial proximity predicts concordance in the direction and magnitude of complex-trait effects has been unclear. We defined effect concordance (*ρ*) as the ratio between mean pairwise causal-effect covariance and mean effect-size variance among selected variant pairs. We stratified variant pairs by three dimensional distance in AlphaFold-predicted structures^18,58^ or by one dimensional coding-sequence distance, and estimated concordance using KASM Baseline + Indep. augmented with kernels encoding these pairs (**Methods**).

Across missense variant pairs, average concordance was *ρ*=0.34±0.01, consistent with a high degree of effect-size heterogeneity. 3D-proximal pairs (<6Å) had greater effect concordance than 3D-distant pairs (≥6Å; *ρ*=0.57±0.04 vs. 0.34±0.02; **Figure 6** and **Supplementary Table 13**). This distance threshold roughly identifies residues that are close enough to make structural contact. Stratified by 1D distance, concordance was *ρ=*0.50±0.03 for pairs separated by <15 nucleotides (nt) versus 0.33±0.01 for pairs separated by ≥90nt. As expected, pLoF variants showed high effect concordance across distance strata (**Figure 6** and **Supplementary Table 13**).

**Figure 6.**
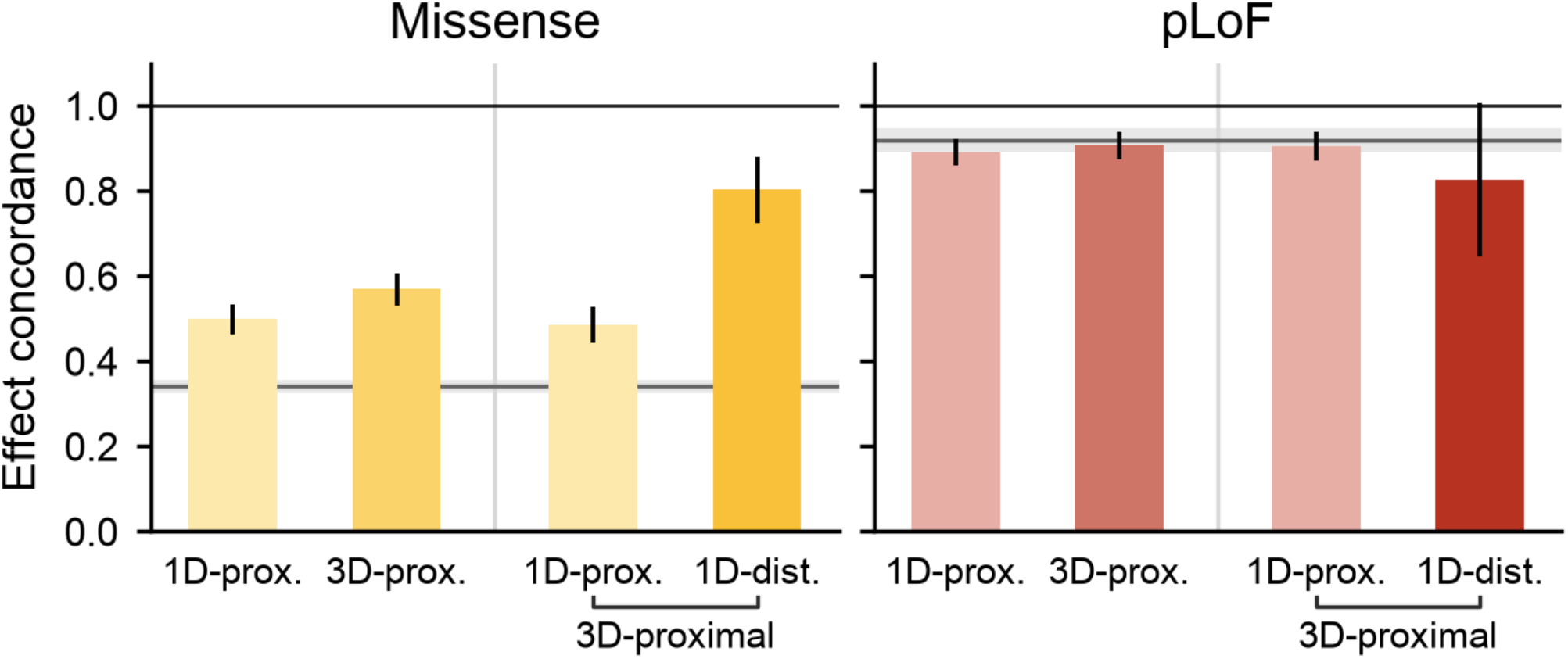
Effect concordance of rare coding variants. Effect concordance (ρ) for within-gene missense and pLoF variant pairs proximal in coding sequence (1D-prox.; <15 nt) or in AlphaFold-predicted structure (3D-prox.; side-chain distance <6 Å), and, among 3D-proximal pairs, for 1D-proximal and 1D-distant (1D-dist.; ≥90 nt) pairs. Gray lines and bands show concordance ± standard error across all within-gene pairs of the same class. Estimates are meta-analyzed across UK Biobank and All of Us traits. Error bars show standard errors.

Notably, among 3D-proximal missense pairs (<6Å), variants that were distant in 1D (≥90nt; 1D-distant) had greater effect concordance than pairs that were also proximal in 1D (<15nt; 1D-proximal; *ρ*=0.80±0.08 vs. 0.48±0.04; *P*=9×10^−5^; **Figure 6, Supplementary Figure 18**, and **Supplementary Table 13**). These 3D-proximal/1D-distant residue pairs are folded into close proximity and may interact to stabilize the fold, such that changing either residue disrupts the interaction or destabilizes the folded structure. Indeed, residues in 3D contact exhibit co-evolution across homologs, supporting the notion that their pairwise interaction determines their substitution effects^59^. We also observed elevated effect concordance among variants within the same secondary-structure element and between hydrogen-bonded residues (**Supplementary Figure 19**). Concordance was higher among missense variant pairs in which both variants had high AlphaMissense scores, consistent with more concordant trait effects among variants predicted to be deleterious (**Supplementary Figure 20**).

We next estimated effect concordance without any distance threshold among missense variants (*ρ*_MM_), among pLoF variants (*ρ*_LL_), and between missense and pLoF variants in the same genes (*ρ*_ML_), restricting to genes with at least two variants of each class (**Methods**). Concordance was substantially higher among pLoF variants (*ρ*_LL_=0.91±0.03) than among missense variants (*ρ*_MM_=0.38±0.02), whereas cross-class concordance was *ρ*_ML_=0.40±0.02. Their normalized ratio 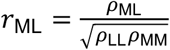was 0.68±0.03, consistent with the previously reported burden genetic correlation^9^.Under a simple partial-loss-of-function model with a shared latent effect direction, *r*_ML_ would equal one, even if missense effects are weaker and more heterogeneous. The lower observed correlation could reflect gain- or change-of-function mechanisms, or gene-to-gene variation in the relative effects of missense and pLoF variants (**Discussion**).

### Characterizing functional allelic series

Functional enrichments may vary across the allelic series of different genes, depending on their particular structure and function. We applied KASM to each of 299 significant gene-trait associations individually and tested for enrichment of 20 structural and functional annotations. We identified 215 significant gene-trait-annotation triplets spanning 79 gene-trait pairs and 19 annotations (one-sided Wald tests of enrichment greater than one; Bonferroni-corrected; **Supplementary Table 14; Methods**). We examined the two largest enrichments, in *PIEZO1* and *FGFR3*, in detail using protein structures and functional assays.

Within *PIEZO1*, a trimeric ion channel associated with HbA1c^60,61^, we identified a striking enrichment of HbA1c heritability at hydrogen-bond contacts connecting individual subunits of the channel (**Figure 7** and **Supplementary Table 15**). These contacts occur at two inter-subunit interfaces, the cytoplasmic anchor and the extracellular cap, where residues form hydrogen bonds with neighboring subunits^60,61^. They include only 0.6% of *PIEZO1* missense variants (14 of 2,472; 0.8% of heterozygosity) yet explain 78% of the gene’s HbA1c heritability (95±3-fold enrichment,*P* = 5×10^−207^, burden Z=−27). 7 of 14 hydrogen-bond contact variants had nominally significant effects, all lowering HbA1c (Z<−3). They map to five positions spanning both interfaces: R2110, T2127 and K2169 in the cytoplasmic anchor, and L2277 and R2279 in the extracellular cap (the latter two are paired, forming a side chain-main chain hydrogen bond). By contrast, heterozygous pLoF variants had weaker effects in the opposite, HbA1c-increasing direction (burden Z = +3.1, *P* = 1.8×10^−3^). These results are consistent with a gain-of-function mechanism in which weaker inter-subunit coupling slows channel inactivation, causing increased cation influx. This influx would cause red blood cell dehydration^62–64^, shorten red cell lifespan, and lower HbA1c^65^. Monti et al.^66^ previously highlighted this association in a functionally informed analysis but did not identify the enrichment at hydrogen-bond residues.

**Figure 7.**
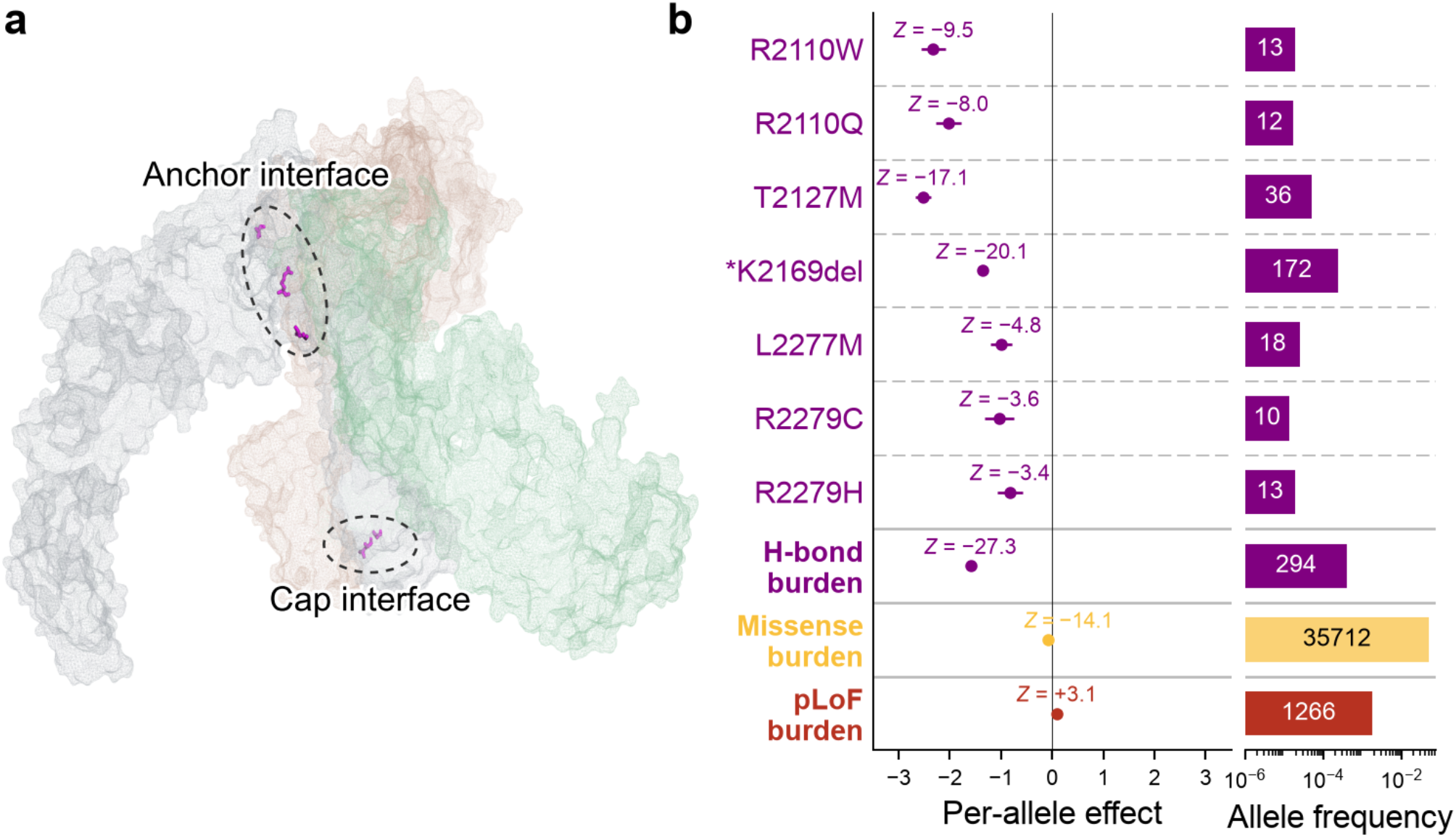
*PIEZO1*-HbA1c allelic series at interchain hydrogen-bond residues. **(a)** Structure of the *PIEZO1* homotrimer (PDB: 8YEZ^85^), with chains shown as mesh surfaces and residues of the interchain hydrogen-bond (H-bond) variants in **(b)** as sticks. Dashed circles mark the cytoplasmic anchor interface (R2110, T2127, K2169) and the extracellular cap interface (L2277, R2279). **(b)** Per-allele HbA1c effects of H-bond variants with |Z| ≥ 3 and burden effects of all H-bond, missense and pLoF variants in UK Biobank, with allele frequencies and allele counts (in bars). *K2169del removes a residue near hydrogen-bonding positions, likely disrupting inter-subunit hydrogen bonds. Error bars show standard errors.

Within *FGFR3*, a transmembrane receptor that regulates skeletal growth^67^ (**Figure 8a**), we identified strong enrichment of height heritability in the transmembrane domain. This domain contains only 3.0% of *FGFR3* missense variants (16 of 524) but explains 44% of the gene’s missense height heritability (9.2±0.8-fold enrichment, *P* = 7.5×10^−27^; **Supplementary Table 16**). The enrichment was driven primarily by G380R, the canonical achondroplasia-associated substitution in the FGFR3 transmembrane helix^68^, which promotes aberrant activation of the FGFR3 receptor tyrosine kinase^69^. We therefore tested whether the genetic effects on height tracked experimentally measured FGFR3 activation using a kinase-domain deep mutational scanning assay^51^, stratifying variants by their FGFR3 activation scores. Variants in the highest-activation-scoring quartile predominantly had negative effect estimates on height (33 of 39 variants; **Figure 8b**) and were strongly enriched for height heritability relative to all *FGFR3* missense variants (5.3±0.5-fold enrichment, *P* = 9.8×10^−17^; **Figure 8c**), whereas the lower-scoring quartiles showed no significant enrichment. This relationship was consistent across experimental conditions (**Supplementary Figure 21**). This example shows how missense associations in different protein domains, here in conjunction with DMS data, can provide convergent mechanistic evidence.

**Figure 8.**
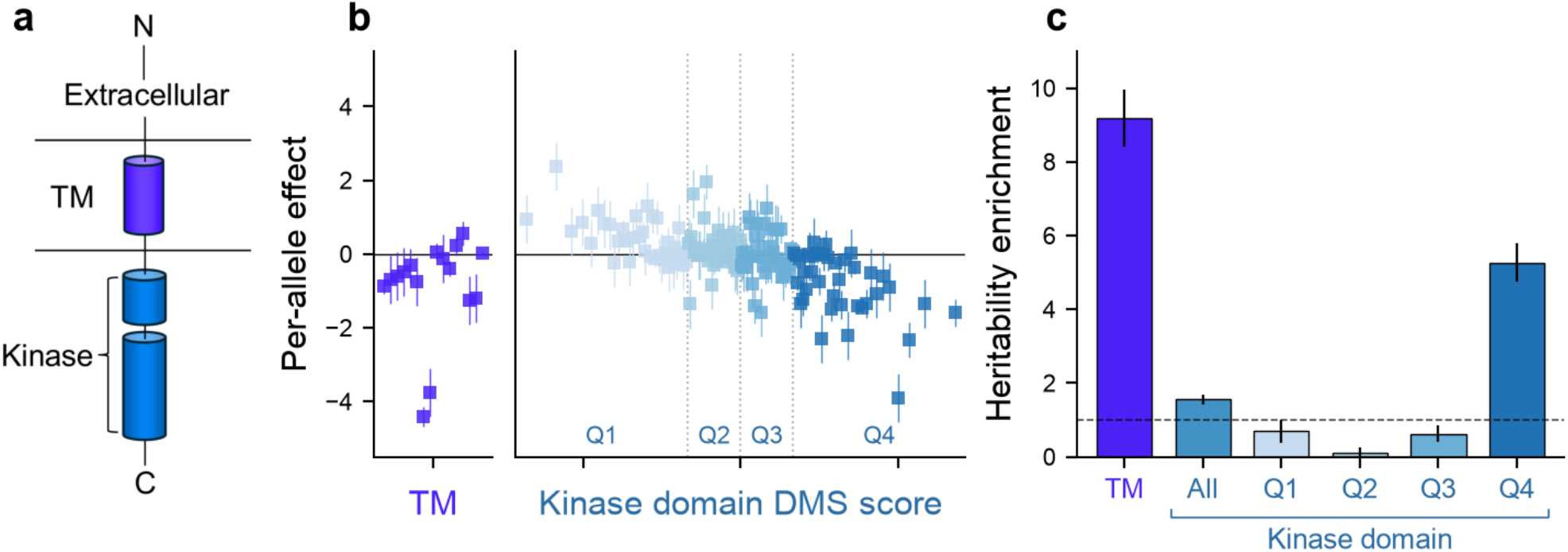
*FGFR3*-height allelic series across transmembrane and kinase domains. **(a)** Schematic of FGFR3 showing extracellular, transmembrane (TM) and kinase domains. **(b)** Per-allele height effects of *FGFR3* missense variants in the TM domain, ordered by residue position, and in the kinase domain, ordered by deep mutational scanning (DMS) activation score and divided into quartiles (Q1-Q4). **(c)** Height heritability enrichment for TM variants, all DMS-scored kinase-domain variants and each DMS quartile. Error bars show standard errors.

We highlight two additional examples involving ion-binding sites (**Supplementary Figure 22**). *CASR*, encoding the Ca^2+^-sensing receptor (CaSR), regulates calcium homeostasis^70^ and is implicated in Mendelian disorders of both increased and decreased serum calcium through loss- and gain-of-function variants^71,72^. In our analysis, missense effects were enriched at Ca^2+^-binding residues: all five missense variants, mapping to four such residues, were associated with higher serum calcium levels, with effect sizes similar to pLoF variants (approximately 2 s.d.). These residues either coordinate Ca^2+^ or couple Ca^2+^ binding to receptor activation^73,74^. The direction of the effect is therefore consistent with partial loss of function that reduces CaSR sensitivity to extracellular Ca^2+^, raising the serum calcium set point^70^. *IHH*, encoding Indian hedgehog, a signaling protein required for growth-plate development^75^, was associated with height, with missense effects enriched at its conserved Zn^2+^-binding site. The two observed H145 variants (H145R and H145N) affect a residue that directly coordinates Zn^2+^ within the IHH signaling domain^76^, and both were associated with shorter stature, also with effect sizes similar to pLoF variants (approximately −2 s.d.). A mutation at the homologous zinc-coordinating histidine in *SHH* was previously shown to substantially reduce zinc affinity, destabilize the folded signaling domain, and diminish signaling activity^77^. The H145 substitutions in *IHH* may similarly impair the stability and activity of the IHH signaling domain. These examples illustrate how integrating structural and functional features adds mechanistic resolution to pLoF burden associations: in both cases, missense alleles at specific functional sites recapitulate the pLoF effect and help to explain what trait-relevant function is lost.

## Discussion

Rare missense variants perturb proteins through diverse mechanisms, producing phenotypic effects that differ in magnitude and direction within the same gene. In clinical genetics, distinguishing pathogenic from benign mutations has been a longstanding goal, and recent progress has been driven by the development of machine learning algorithms for variant effect prediction^10–17^. These algorithms, in turn, were enabled by fundamental advances in algorithms to predict protein structure and function. We combined deleteriousness predictors with a range of functional features, many of them predicted computationally, to provide molecular explanations for how protein-altering variants affect human complex traits. With this approach, missense variants contributed more heritability than pLoF variants, their effects were concentrated in structurally and functionally important protein features, and mutations at 3D-proximal residues had highly concordant phenotypic effects. Within individual genes, enriched annotations pointed to plausible mechanisms. KASM therefore connects allelic heterogeneity to protein structure and function both exome-wide and within individual gene-trait associations.

A previous study^9^ estimated a missense burden heritability of around 0.5% on average across traits, providing a lower bound on the total contribution of rare missense variants. KASM substantially improves this lower bound, from 0.54±0.04% under a BHR-like burden model to 1.27±0.09% under KASM Baseline x Exon and 1.83±0.11% for KASM with independent variant effects. The latter estimate is about 2x higher for missense versus pLoF variants.

These estimates were also larger than those reported in a recent whole-genome sequencing analysis in UK Biobank, which analyzed variants with MAF > 0.01%^36^. For height, that study estimated that coding variants with MAF of 0.01-1% explained 3.5% of phenotypic variance in height. We estimated that missense and pLoF variants with MAF < 0.001%, 0.001-0.01% and 0.01-0.1% explained 5.5%, 3.0%, and 1.9% of phenotypic variance in height, respectively; our estimate in the overlapping 0.01-0.1% range was consistent with the published estimate of 3.5% in the broader 0.01-1% range. These results suggest that rare coding variants account for a substantial fraction of heritability not captured by previous analyses. For height, our estimate for variants with MAF < 0.01% corresponds to approximately half the gap between WGS-based heritability of 70.9% and the reported pedigree-based heritability of 88.2%^36^.

Variant-effect predictors are commonly evaluated against clinical or experimental benchmarks that may not directly measure their ability to prioritize variants affecting complex traits^78–80^. KASM asks whether a predictor identifies variants with disproportionately large effects on complex traits. Unlike clinical pathogenicity benchmarks, which are concentrated in established Mendelian disease genes, KASM evaluates variants across the broader range of genes and traits in population-scale exome studies. It also complements DMS assays, whose molecular readouts are measured in specific experimental contexts and may not translate directly to organism-level phenotypes or disease risk. In our comparisons, computational predictors sometimes stratified trait effects more strongly than DMS, indicating that DMS performance is not necessarily gold-standard evidence for variant prioritization.

On the other hand, general-purpose deleteriousness predictors such as AlphaMissense do not provide mechanistic hypotheses, and interpretable functional annotations such as DMS scores complement such predictors by helping identify the molecular function of deleterious alleles. At *FGFR3*, variant effects on height tracked experimentally measured kinase activation, supporting altered receptor signaling as a mechanism linking missense variation to the phenotype. At *PIEZO1*, HbA1c heritability was concentrated among missense substitutions at inter-subunit hydrogen-bond contacts, supporting a mechanism involving gain of function. Exome-wide analyses demonstrate the same principle: for example, charge-reversing substitutions at interchain hydrogen bond residues have disproportionately large effects, implying that an important coding-variant mechanism is the disruption of quaternary structure.

Several existing methods integrate functional annotations to improve the power of rare variant association tests. Monti et al.^66^ use functionally informed kernels to increase gene-level association power, motivated as we are by the principle that variants with similar functional annotations might have similar phenotypic effects, but a key difference is that our kernels are estimated from the data, while theirs are fixed. STAAR^81^ uses a similar approach, combining the P values of multiple SKAT and annotation-weighted burden tests. DeepRVAT^27^ does learn a data-dependent variant aggregation function, but the output of this function is a single score per gene, leading to a burden-like rather than kernel-like association test. Each of these approaches overlaps with ours methodologically, but their goal is different.

An unresolved question is how often missense variants have effects other than partial loss of function. For rare diseases, a substantial fraction of known causal alleles acts via gain of function^82^. We found that for complex traits, missense and pLoF effects in the same gene do have a positive correlation, but this correlation is weaker than expected under a narrow model of uniformly partial loss-of-function effects. Within pLoF-constrained genes, heritability enrichment is similar for pLoF alleles and AlphaMissense-prioritized missense alleles, consistent with those alleles causing partial loss-of-function; on the other hand, constrained genes could be similarly sensitive to both loss and gain of function^83^. Overall, we can rule out a narrow model in which missense variants produce weaker loss-of-function effects in a manner that is uniform across genes, but addressing the broader question remains challenging because existing annotations do not systematically identify gain-of-function effects.

More generally, a key limitation of KASM is that it relies upon existing annotations, which might not capture the functional perturbations through which variants influence traits. More accurate deleteriousness predictors^14,17^ could yield higher heritability estimates, and annotations of specific molecular mechanisms, including ligand/protein binding^21–23^ and post-translational regulation^5,84^, could provide new explanations for how variants influence traits.

We note several additional limitations. First, KASM fits an additive model among component kernels, without interactions except involving frequency bins. This approach is not expected to produce bias, but it potentially misses interesting interactions, and indeed, certain combinations of annotations do exhibit notable enrichments (**Figure 5**). Second, our exome-wide analyses provide limited insight into heterogeneity among genes; for example, if some annotations were enriched specifically in some gene family, this enrichment would be diluted in our analysis. We do show that significant gene-trait pairs sometimes exhibit strong enrichments, pointing to gene-specific biology. Third, our analysis is limited to protein coding variants; regulatory variants would pose additional methodological challenges, including the problem of how to map variants to genes or otherwise group them into units. Despite these limitations, our results provide insight into the molecular processes through which protein variation influences human phenotypes.

## Methods

### Functionally informed kernels

We model rare coding effects using functionally informed kernels that encode assumptions about the distribution of variant effect sizes within genes. Let **y** ∈ ℝ^*N*^ denote a standardized phenotype vector, with mean 0 and variance 1 across *N* individuals. Let 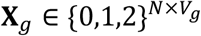denote the genotype dosage matrix for the *V*_*g*_ rare coding variants (minor allele frequency (MAF)<10^−3^) for gene *g* = 1, …, *G*. Concatenating across genes, define **X** = [**X**_1_, …, **X**_G_] and 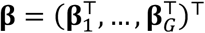. The linear model is

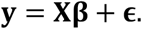

We assume that effect sizes are independent across genes but that variant effects are structured within each gene. Specifically, the per-allele effect size vector **β**_*g*_ for each gene *g* has mean zero, and covariance given by a gene-scaled combination of functionally informed kernels:

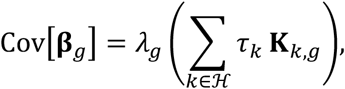

where ℋ indexes the heritable kernels, 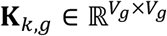 is the *k*^th^ heritable kernel for gene *g, τ*_*k*_ is its coefficient shared across genes, and *λ*_*g*_ is a gene-specific scaling factor used to stratify genes by selective constraint. We estimate {*τ*_*k*_} and {*λ*_*g*_} in a manner that accounts for within-gene linkage disequilibrium (LD) and overdispersion in exome-wide association summary statistics. The heritable kernels can take various forms, as described below.

#### Burden kernels

To model effects that share a common direction among variants within a gene (with sign varying across genes), we use burden kernels. Let 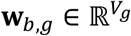 denote the *b*-th burden weight vector for gene *g*. Its entries can be binary, such as indicators of damaging missense variants, or continuous, such as a deleteriousness score. The corresponding rank-one kernel is

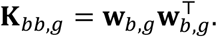

This kernel assigns each pair of variants an effect size covariance proportional to the product of their weights, thereby modeling stronger shared effects among variants with larger weights along the burden direction.

#### Cross-burden kernels

To model covariance between two distinct burden directions, we use cross-burden kernels. For two burden-weight vectors **w**_*b,g*_ and **w**_*b,g*_, the symmetric cross-burden kernel is

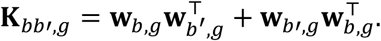

This kernel is relevant both to pairs of variants and to individual variants. For pairs of variants, it allows the effects of variants prioritized by one burden weight to covary with the effects of variants prioritized by another burden weight, such as missense and pLoF variants within the same gene. For individual variants, it affects the effect-size variance of variants in the intersection of two burden weight annotations.

#### MAF-stratified burden kernels

To allow burden effect covariance to vary by MAF, we stratify each burden weight by allele frequency bin. Let 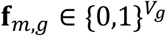 denote the mask for frequency bin *m*, with entries equal to 1 for variants in the bin and 0 otherwise. The masked burden weight is

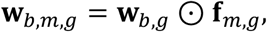

where ⊙ denotes element-wise multiplication. We construct burden and cross-burden kernels from **w**_*b,m,g*_ as defined above, allowing the same functional annotation to contribute differently across allele frequency bins.

#### Position kernels

To model covariance between proximal variants, we define a binary position kernel 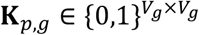 for each gene *g*. Its entries indicate whether each pair of distinct variants satisfied a specified proximity criterion (see below).

#### Gene-level scaling

To allow the magnitude of rare coding effects to vary across genes—reflecting both how strongly a variant perturbs a gene and how strongly that gene influences the phenotype—we scale all heritable kernels for gene *g* by a gene-specific factor *λ*_*g*_. We parametrize *λ*_*g*_ using five bins of gnomAD gene-level pLoF observed-to-expected ratio (constraint quintile). Let *Z*_*g*_ ∈ {1,2,3,4,5} denote the constraint bin for gene *g*, and define 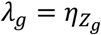, where *η*_z_ ≥ 0 is the scaling coefficient for bin *z*. To ensure identifiability between the shared kernel coefficients {*τ*_*k*_}_*k*∈ℋ_ and the gene-level scaling coefficients **η**, we impose ^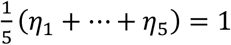^.

### Kernel estimands

We next define estimands associated with a prespecified set of functionally informed kernels.

#### Kernel similarity and the best-fitting kernel

Let **X**_*i*_ denote the centered genotype vector for individual *i*, let 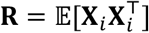 denote the genotype covariance matrix, and let **β** be a random vector of effect sizes. KASM features both a true effect-size covariance matrix, **Σ** = E[**ββ**^⊤^], and a prespecified space of possible kernels, *K*, which may or may not contain **Σ**. In this work, *K* is the linear span of the functionally informed kernels. For individuals *i* ≠ *j*, define the genetic similarity induced by a kernel **A** as 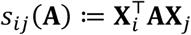. Two kernels **A** and **B** are similar if they induce similar genetic similarity functions: specifically, we define a matrix inner product

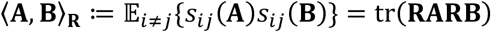

with corresponding norm 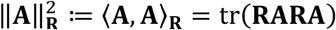. This construction quantifies the similarity between the true kernel **Σ** and a member **K** of *K*; the squared correlation between the genetic similarities induced by **K** and **Σ** is:

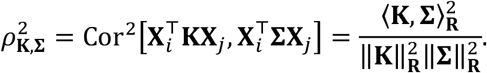

Let *P*_*K*_ denote orthogonal projection onto *K* under the inner product ⟨⋅,⋅⟩_**R**_. The best approximation to the true effect-size covariance within the kernel space is therefore

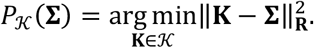

Equivalently, this projection maximizes the squared correlation between the genetic similarities induced by **K** and **Σ** over *K*. We refer to **K**\* = *P*_*K*_ (**Σ**) as the best-fitting kernel.

#### Kernel heritability

We define kernel heritability as the SNP-heritability explained by the best-fitting member of a prespecified set of kernels. By convention, we use the term “SNP-heritability” to denote the proportion of phenotypic variance explained by the causal and LD-tagging effects of a specified set of variants, which may include both single-nucleotide variants and insertions or deletions.

The additive genetic component of the phenotype is 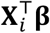, and the total heritability tagged by the measured variants is

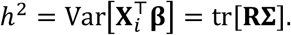

We define the kernel heritability as the genetic variance corresponding to the projected effect-size covariance:

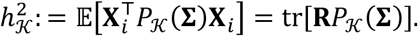

#### When the kernel heritability is a lower bound

Noting that tr(**RA**) = ⟨**A, R**^−1^⟩_**R**_ for every symmetric **A**, and because *P*_*K*_ is self-adjoint under ⟨⋅,⋅⟩_**R**_,

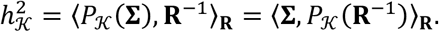

Similarly, the SNP-heritability is *h*^2^ = ⟨**Σ, R**^−1^⟩_**R**_. These identities characterize when kernel heritability is bounded above by the total SNP-heritability. We write **A** ≽ **B** to mean that the matrix **A** − **B** is positive semidefinite. For any symmetric matrix **A**, ⟨**Σ, A**⟩_**R**_ ≥ 0 for every **Σ** ≽ 0 if and only if **A** ≽ 0. Consequently, the necessary and sufficient condition to ensure

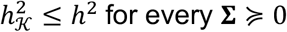

is that

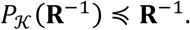

One sufficient condition for this to hold is that *K* has the form *K*_**U**_ = {**UTU** ^⊤^: **T** = **T** ^⊤^}, which is the case for a single-gene burden kernel model. Another sufficient condition is that **R**^−1^ ∈ *K*, in which case *P*_*K*_(**R**^−1^) = **R**^−1^ and the kernel heritability equals the total heritability. We added **R**^−1^ as an additional overdispersion kernel alongside the three MAF-stratified kernels. Heritability estimates were nearly unchanged across the baseline, exon-augmented, and independent-effect models, and the overdispersion component was nonnegative, indicating that the kernel heritability is a lower bound for these models (**Supplementary Figure 14**).

#### Variance components and overdispersion kernels

The kernel estimand extends to several jointly fitted components. Suppose that the kernel space is a direct sum *K* = *K*_1_ ⊕ ⋯ ⊕ *K*_*Q*_, so that every fitted kernel has a unique decomposition into elements of the component spaces:

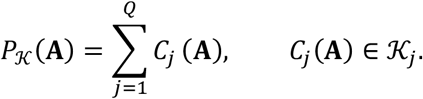

Each term makes a contribution to phenotypic variance:

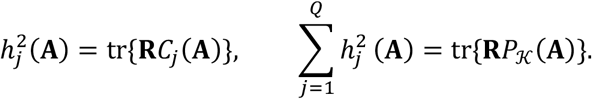

In particular, let *K* = *K*_*o*_ ⊕ *K*_*h*_ decompose the kernel space into overdispersion and heritable kernel spaces. Writing *P*_*K*_ (**A**) = *C*_*o*_(**A**) + *C*_*h*_(**A**), we define the heritable-component estimand as

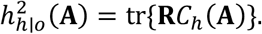

This estimand depends on both kernel spaces. Let 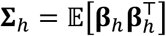 and 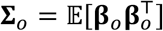 denote the biological and confounding effect second moments, respectively; **Σ**_*h*_ is the biological matrix **Σ** defined above. Assuming zero cross second moments between these effects, their combined second moment is **Σ**_*h*+o_ = **Σ**_*h*_ + **Σ**_*0*_. If

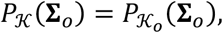

then the projection of the confounding effects lies entirely in the overdispersion space, so *C*_*h*_(**Σ**_B_) = 0. The component maps are linear, and therefore

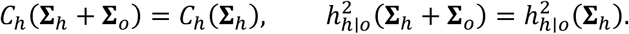

This condition is weaker than **Σ**_B_ ∈ *K*_B_; it permits covariance structure among the overdispersion effects which is not captured by any overdispersion kernel, so long as this structure is also not captured by any heritable kernel. For example, population stratification effects might have genome-wide low-rank structure; the assumption would be that a functional annotation has no signed correlation with variant loadings. Weiner and Nadig et al.^9^ quantified a plausible form of stratification, minor allele-biased stratification, which would violate this assumption (see below).

#### Special cases of the kernel heritability

We consider several examples of the kernel space *K* and its corresponding kernel heritability.

- If the true effect-size covariance lies in the kernel space, **Σ** ∈ *K*, then *P*_*K*_(**Σ**) = **Σ**, so the estimand recovers total heritability 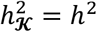.
- If the kernel space is the linear span of a single kernel **K**, that is, *K* ≔ {τ**K**, τ ∈ ℝ}, then 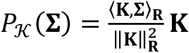, and the corresponding kernel heritability is 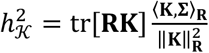. In particular,if **K** = **R**^−1^, then 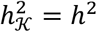
- If the kernel space is generated by a rank-one burden kernel with burden weight vector **w**, *K* = {*τ***ww** ^⊤^: *τ* ∈ ℝ}, then the kernel heritability is the variance explained by the corresponding burden score, 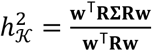.
- For the heritable component estimand, take *K*_C_ = span{**J**} and *K*_B_ = span{**I**}, where **J** = **11** ^⊤^, and assume no LD and at least two variants. Under the BHR model, per-allele effects have the form **β** = *µ***1** + **δ**, where *E*[**δ**] = 0 and Cov(**δ**) = **D** is diagonal, giving **Σ** = *µ*^2^**J** + **D**. Residualizing **J** against **I** gives **J** − **I**, which is orthogonal to every diagonal matrix under ⟨⋅,⋅⟩_**R**_ when **R** is diagonal. It follows that *C*_*h*_(**Σ**) = *µ*^2^**J**, which means that the burden kernel absorbs the burden effect size *µ* while the overdispersion kernel absorbs the overdispersion effects (biological or otherwise). The corresponding kernel heritability is 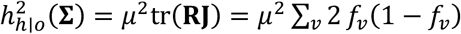, where *f*_*v*_ is the allele frequency. This agrees with the burden heritability definition of Weiner and Nadig et al.

#### Heritability enrichment

To quantify whether variants in an annotated set contribute disproportionately to heritability, we compare their fraction of kernel heritability to their fraction of total rare-variant heterozygosity. Let *A* denote an annotated variant set, **K** = *C*_*h*_(**Σ**), the fitted heritable effect-size covariance, and **R** the LD matrix. The heritability represented by **K** is *h*^2^(**K**) = tr(**RK**). When the off-diagonal contributions *R*_*vv*,_*K*_*vv*_ average to zero across variant pairs, this quantity reduces to *h*^2^(**K**) = ∑_*v*_ *R*_*vv*_*K*_*vv*_, where *R*_*vv*_ is the heterozygosity of variant *v* and *K*_KK_ is its effect size variance. The heritability attributable to *A* is therefore 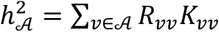. We define heritability enrichment as:

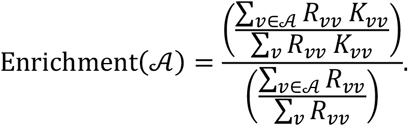

If variants in annotation *A* have the same expected per-allele squared effect size as all variants, the enrichment equals 1; values above 1 indicate larger per-allele squared effects.

#### Effect concordance for variant pairs

To quantify effect concordance among prespecified pairs of variants, let 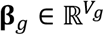 denote the vector of per-allele causal effects in gene *g*, and let *P*_*g*_ denote a set of variant pairs from the same or different functional classes. We define concordance as the mean effect-size cross-product divided by the geometric mean of the two paired mean squared effects:

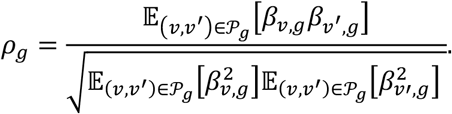

We aggregate across genes by summing the pairwise covariance numerator and the two variance denominators separately and then taking their ratio:

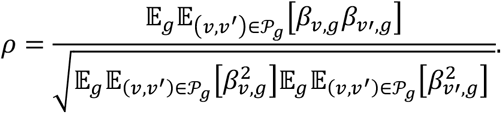

If every selected pair has identical nonzero effects, *ρ*=1. If paired effects are independent with mean zero, the expected cross-products and hence *ρ* are zero. We use the following pair sets. For positional concordance, *P*_g_ contains distinct variant pairs satisfying a prespecified 1D or 3D distance criterion.

For within-class concordance, *P*_g_ contains distinct pairs from the same functional class; for cross-class concordance, *P*_g_ contains pairs with one missense and one pLoF variant. We restrict these analyses to the same set of genes containing at least two variants of each class. We denote within-missense concordance by *ρ*_MM_, within-pLoF concordance by *ρ*_LL_, and missense-pLoF concordance by *ρ*_ML_. We further define 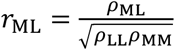 which gives a deattenuated correlation between the underlying gene-level missense and pLoF burden effects. Equivalently, under a model in which individual variant effects fluctuate around class-specific mean burden effects within each gene, *r*_ML_ corresponds to the Pearson correlation between those mean missense and pLoF burden effects across genes.

### Inference for the Kernel Allelic Series Model

KASM operates on variant association statistics, variant-level functional annotations and gene-level constraint bins, and within-gene LD matrices. It applies separate random-effects and fixed-effects estimators for subthreshold and exome-wide significant gene-trait associations. The random-effects estimator models covariance shared across subthreshold genes and includes overdispersion components for residual inflation. The fixed-effects estimator conditions on each significant gene’s realized effects without imposing a distributional assumption. Compared with the random-effects model, an advantage of the fixed-effects estimator is that it provides gene-specific estimates; a disadvantage is that it does not include a correction for confounding. The rationale for combining both estimators is that excluding significant genes stabilizes the random-effects estimator, while excluding subthreshold genes limits the effect of confounding on the fixed-effects estimator. The same separation is used by BHR^9^ and LDSC^86^; a difference is that our fixed-effects estimator is more sophisticated.

### Random-effects estimator for subthreshold genes

For subthreshold genes, we distinguish biological effects 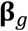 from population effects 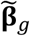, where the latter also includes non-biological contributions from population stratification and cryptic relatedness. Dynastic effects and effects due to assortative mating would be included in **β**_*g*_; more generally, any effects which are mediated by changes to the protein, and thus might align with functional annotations, are included. We specify a kernel space for 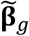 involving heritable kernels, which are interpreted as biological, and overdispersion kernels, which are interpreted as non-biological:

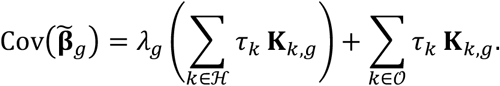

ℋ indexes the heritable kernels, *O* indexes the overdispersion kernels, and *λ*_*g*_ is the gene-level scaling factor. The overdispersion kernels capture residual confounding and are not multiplied by *λ*_*g*_. We use diagonal overdispersion kernels, **K** _*k,g*_ = diag(**f**_*k,g*_), *k* ∈ *O*, where **f**_*k,g*_ is the allele frequency bin mask for overdispersion component *k*.

With 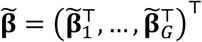, the phenotype model is

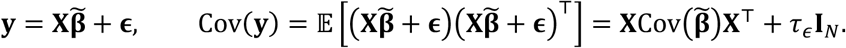

The unknown parameters are the heritable kernel coefficients {*τ*_*k*_}_*k*∈ ℋ_, the gene-scaling coefficients **η**, the overdispersion coefficients {*τ*_*k*_}_+∈*O*_, and the residual parameter *τ*_*ϵ*_.

We estimate gene-level parameters ***η*** and kernel-level parameters ***τ*** by alternating updates. Each update holds one parameter set fixed, represents the covariance as a linear combination of prespecified kernels, and solves a Haseman-Elston regression.

#### Kernel-coefficient update

With **η** fixed, and hence *λ*_*g*_ known,

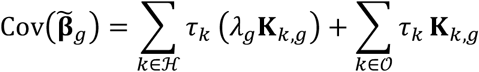

which is linear in the heritable-kernel and overdispersion coefficients {*τ*_*k*_}_*k*+∈ℋ_, {*τ*_*k*_}_*k*∈*O*_.

#### Gene-coefficient update

With {*τ*_*k*_}_*k*+∈ℋ_ fixed, define the aggregate heritable kernel **K** _*τ,g*_ = ∑_*k*∈ ℋ_ *τ*_*k*_ **K**_*k,g*_. Then

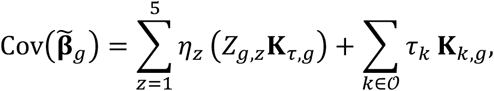

where *Z*_*g,z*_ indicates whether gene *g* is in constraint bin *z*, which is linear in the gene-scaling and overdispersion coefficients **η**, {*τ*_*k*_}_*k*∈*O*_.

#### Haseman-Elston regression

In either update, the covariance is a linear combination of prespecified kernels. Let ℐ denote the kernels in the current update and **K**_*k*_ = blockdiag (**K**_*k*,1_, …, **K**_*k,G*_); then the model being fit is:

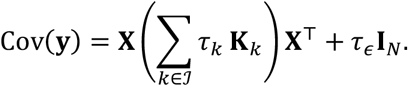

We estimate **τ** and *τ*_*ϵ*_ by minimizing

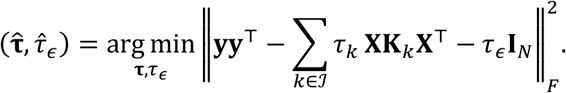

This yields the normal equations

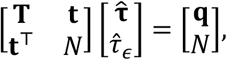

where **T** ∈ ℝ^|ℐ|×| ℐ|^ and **t, q** ∈ ℝ^| ℐ |^, with entries

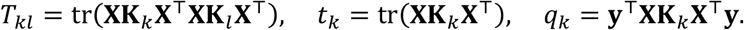

#### Summary-statistic implementation

The normal equation quantities can be evaluated from within-gene LD matrices and variant association summary statistics, without individual-level genotypes or phenotypes. For gene *g*, let 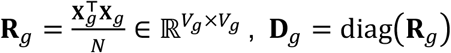 and 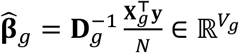. Writing 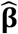, **R** and **D** for the corresponding genome-wide vector and block-diagonal matrices, the normal-equation entries are

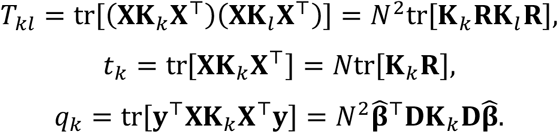

The heritable-component estimate is 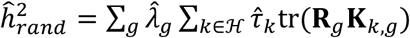. Uncertainty is estimated by a block jackknife over genes. We partition genes into *n*_*jk*_ = 100 blocks with approximately equal numbers of genes. For each block, we subtract its contribution from the genome-wide sufficient statistics, refit the complete alternating system, and compute a leave-one-block-out estimate 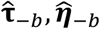. For a scalar estimate *τ*, the jackknife standard error is:

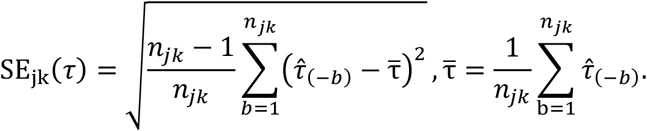

These standard errors represent two sources of uncertainty: sampling noise in the variant association statistics, which would change if the study were repeated in new individuals, and heterogeneity in the genetic effects realized across different parts of the genome.

#### Heritability enrichment estimator

For a binary annotation *A*, we fit KASM Baseline + Annotation (see below) and construct the fitted heritable effect-size covariance kernel 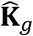 for each gene. We estimate the heritability attributable to *A* and the total heritability as

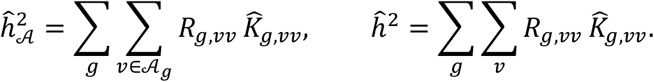

For every leave-one-block-out jackknife estimate, we reconstruct 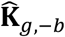 and calculate the enrichment. Its standard error is the block-jackknife standard error across the resulting leave-one-block-out ratios.

#### Relationship to burden heritability regression (BHR)

In a simplified setting, the KASM estimator is exactly equivalent to BHR. We consider the case of a single rank-one burden kernel, with no gene-level scaling, no fixed-effect genes, and no overdispersion component. For each gene *g*, let

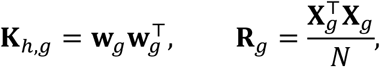

and define the burden genotype, its variance, and its standardized association statistic as

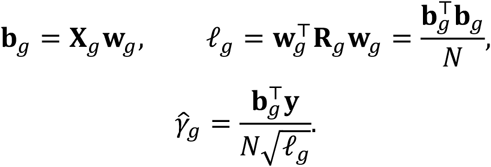

The phenotype covariance model is

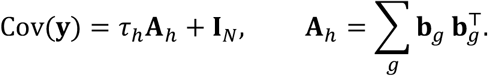

With the residual variance fixed, the one-parameter Haseman–Elston normal equation is

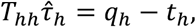

where

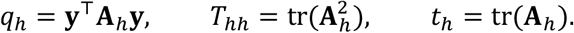

These quantities can be written in terms of the burden association statistics. First,

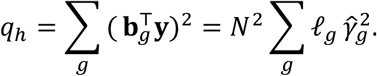

Second, ignoring LD between genes,

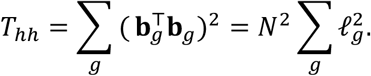

Finally,

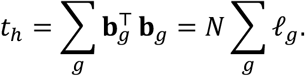

Substitution into the Haseman–Elston normal equation gives

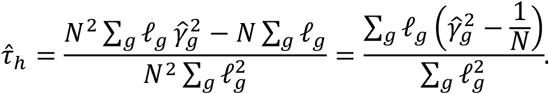

Under the same model,

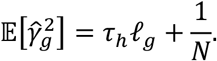

Thus, the KASM Haseman–Elston estimator is exactly the ordinary least-squares BHR estimator obtained by regressing 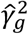 on *l*_*g*_ with the intercept fixed at 1/*N*:

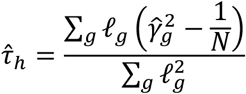

A similar equivalency is known between HE Regression and LD Score Regression^87^.

### Fixed-effects estimator for significant genes

For exome-wide significant genes, we model variant effects within each gene using gene-specific fixed burden effects. For gene *g*, let 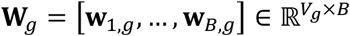 denote burden-weight vectors and let **B**_*g*_= **X**_*g*_**W**_*g*_ ∈ ℝ^*N*×*B*^ denote the corresponding burden-score matrix. We use the fixed-effects model

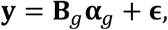

where 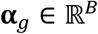 is the vector of gene-specific burden-effect parameters.

#### Relationship to random-effects model

If **α**_*g*_ were instead a mean-zero random vector with shared covariance Cov[**α**_*g*_] = **T** ∈ ℝ^*B* ×*B*^ across genes, the covariance of the gene-level genetic component would be

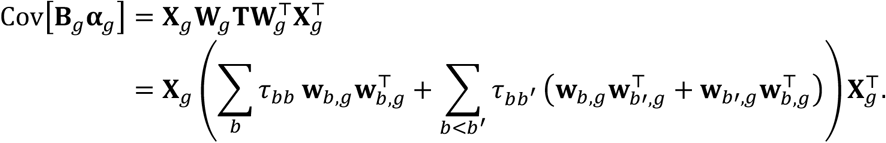

The diagonal entries of **T** correspond to burden kernels, and the off-diagonal entries correspond to cross-burden kernels. Thus, the fixed- and random-effects estimators use the same burden-effect space: the fixed-effects estimator targets the realized effects in each significant gene, whereas the random-effects estimator estimates their shared covariance structure across subthreshold genes.

#### Heritability estimator

Let 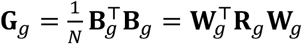 denote the covariance matrix of the burden values. The fixed-effects heritability of gene *g* is:

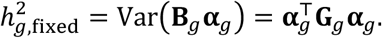

Recall that 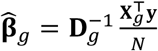, we define the corresponding burden-phenotype covariance vector as 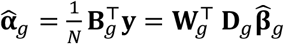. Let *r* = rank(**G**_*g*_), and let 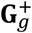denote the matrix pseudoinverse of **G**. An unbiased estimator is

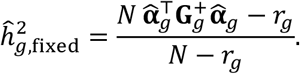

The corresponding approximate sampling standard error is:

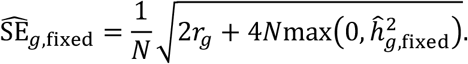

Fixed-effects inference conditions on the analyzed genes and their realized genetic effects. Its standard errors quantify how estimates would vary if association statistics were obtained from a new sample of individuals while the underlying genetic effects remained fixed. Details of this fixed-effects estimator can be found in ref.^44^.

#### Heritability enrichment estimator

For a binary variant annotation *A*, we fit “KASM Baseline + Annotation” (see below) and estimated the annotation heritability 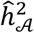 and the total heritability ĥ^2^. We calculated the enrichment and its standard error by the multivariate delta method using the sampling covariance of the annotation and total heritability estimates.

#### Combining random-effects and fixed-effects estimates

We sum fixed-effects heritability over the disjoint set of significant genes 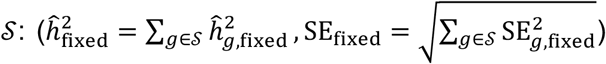 and add it to random-effects heritability from subthreshold genes 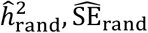. Because the two components use non-overlapping gene sets, the mixed-effects estimate and standard error are:

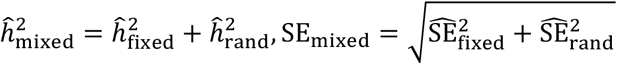

#### Meta-analysis across traits

We combined estimates across traits with fixed, nonnegative weights *w*_*t*_, normalized to sum to one. Estimates in Figure 2 used sample sizes, *w*_*t*_ ∝ *N*_*t*_. For missense heritability enrichment, we used 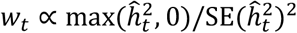, where 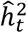 is the KASM Baseline missense heritability. For effect concordance, 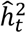 is the KASM Baseline + Indep. heritability of the corresponding variant class, summed over missense and pLoF for cross-class concordance. For ratio estimands (enrichment and concordance), we computed the ratio of the sums of numerators and denominators. For subthreshold genes, we formed each of the 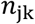 pooled leave-one-block-out replicates as the weighted sum of the corresponding replicates and applied the block-jackknife formula to the pooled replicates. For significant genes, we pooled annotation-specific and total fixed-effects heritability as 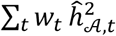and 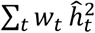, with variances and covariance 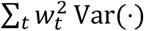 and 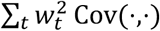. Among 299 significant gene-cohort-trait associations, 260 were distinct gene-cohort combinations and 31 of these appeared in multiple traits, so the omitted cross-trait covariance has a modest effect.

#### Modeling independent effects

To estimate heritability without imposing a burden assumption, we fit a separate fixed effect for every variant in each significant gene. For gene *g*, we replace the burden-score matrix **B**_*g*_ = **X**_*g*_**W**_*g*_with the full genotype matrix **X**_*g*_, which is equivalent to setting 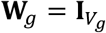 in the above fixed-effects model. Let 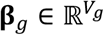 denote the vector of variant-level fixed effects, and the corresponding fixed-effects model is

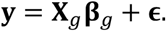

Let *r*_*g*_ = rank (**R**_*g*_), and let 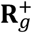 denote the pseudoinverse of **R**_*g*_, the corresponding estimator for gene *g* is

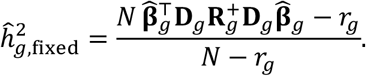

#### Prioritizing significant genes

For each trait and variant class, we identified exome-wide significant genes using an optimized one-dimensional burden test based on the KASM Baseline model. Recall that each column of the burden-score matrix **B**_*g*_ = **X**_*g*_**W**_*g*_ captures a distinct component of variant deleteriousness. We combine these weights using a shared coefficient vector **θ** ∈ ℝ^*B*^, yielding the one-dimensional burden score **b**_*g*_ (**θ**) = **X**_*g*_**W**_*g*_**θ**. The vector **θ** is estimated per trait. This combination leverages the complementary information captured by the individual burden weights while retaining a one-degree-of-freedom test, which can be more powerful than testing all burden weights jointly when their effects align along a shared direction. Recall that 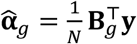 and 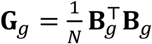. For a given **θ**, the burden-phenotype covariance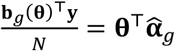, and its sampling variance under the null is Var 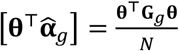. The corresponding chi-square statistic is therefore 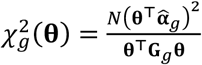, which follows a chi-square distribution with one degree of freedom under the null. Because its noncentrality determines burden-test power, we estimate a shared coefficient vector by maximizing the aggregate chi-square statistic across genes: 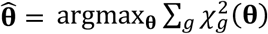. Because **θ** contains only *B* parameters and is estimated jointly across *G* genes,with *G* substantially larger than *B*, this exome-wide optimization limits overfitting and prevents the burden score from being tailored to any single gene. For each gene, we construct the optimized burden score 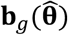 and perform a two-sided burden test using 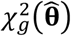. We designate genes with *P* < 0.05/*G* as exome-wide significant, where *G* is the number of tested genes.

### Alignment of stratification effects with functional annotations

Population stratification can bias our random effects estimator, despite the inclusion of overdispersion kernels, if its effects specifically align with functionally informed kernels. In particular, if minor alleles tend to associate with increased trait values, their effects are concordant along the burden direction. Minor allele-biased stratification can also affect BHR, and Weiner and Nadig et al.^9^ showed that it arises in demographic simulations involving subpopulations of different sizes. Unlike biological effects, these stratification effects are expected to be equally correlated for any two minor alleles whether or not they belong to the same gene. Under this assumption, Weiner and Nadig et al. estimated the bias affecting synonymous variant burden heritability in UK Biobank traits to be around 0.01%-0.1% across traits. This calculation involves quantifying the exome-wide burden effect size, as though every variant belonged to a single gene.

For KASM, the relevant form of stratification is slightly more general, since in principle, stratification effects could align specifically with some functional annotation. We therefore generalized the exome-wide burden diagnostic. Specifically, we concatenate variants across genes into one pseudo-gene and apply random-effects KASM Baseline. This approach is informative because any covariance between stratification effects is expected to persist across genes, whereas biological effect covariance is expected to be much larger within genes than across them.

Formally, let 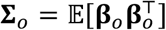, where **β**_*o*_ is a vector of stratification effects, and let *K*_exome_ be a space of whole-exome kernels. Let *K*_gene_ be the space of gene blockwise kernels, obtained by setting to zero the inter-gene entries of each whole-exome kernel **K** ∈ *K*_exome_: its entries have the form **K** ∘ **Δ**, where Δ_*vv*_ is an indicator for *v, v*′ belonging to the same gene. The assumption that stratification effects are the same between and within genes can be expressed as:

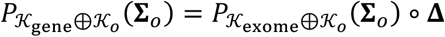

which, assuming zero between-gene LD (**R** ∘ **Δ** = **R**), implies that

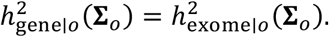

Assuming zero covariance between biological and stratification effects as above, the pooled analysis of observed associations estimates:

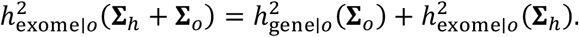

Assuming the second term is nonnegative, this means that the diagnostic gives an upper bound for the inflation term 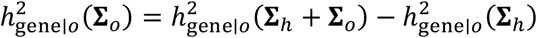.

### Estimating effect concordance

We estimated effect concordance across genes under the “KASM Baseline + Indep. + Position” model (see below). For each gene *g*, let 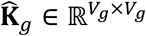 be the causal-effect covariance estimated by the fitted heritable kernels. Let *P*_*g*_ be the set of variant pairs satisfying a given relationship, we estimate the concordance using

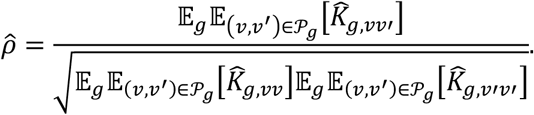

Here, the denominator weights each variant’s effect variance by its number of selected partners within the gene, consistent with the definition of *ρ*.

### Computational techniques

We use several computational techniques to accelerate KASM inference.

#### Sparse-Plus-Low-Rank (SPLR) matrix operations

The random-effects estimator repeatedly evaluates traces of the form tr[**K**_*k*_**RK**_*l*_**R**]. We represent each kernel and LD matrix as an SPLR matrix (**S** + **LQ** ^⊤^), where **S** is sparse and **LQ** ^⊤^ is low rank. Products of SPLR matrices are SPLR:

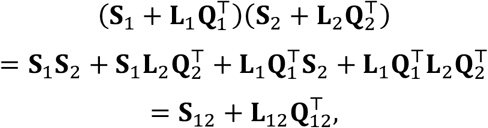

where **S**_12_ = **S**_1_**S**_2_ is a sparse matrix, and if **L**_6_ and **Q**_6_ have *r*_6_ columns, 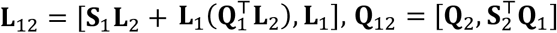 each have *r*_1_ + *r*_2_ columns, and therefore rank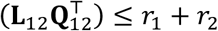 We can consequently evaluate traces using tr(**S** + **LQ** ^⊤^) = tr(**S**) + tr(**Q** ^⊤^ **L**), which requires only the diagonal of the sparse component (∑_6_ *S*_66_) and the trace of the product of the low-rank matrices (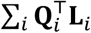 for each column *i*), without constructing dense variant-by-variant matrices.

#### SPLR LD matrices

We compute within-gene LD covariance matrices using individual-level genotype data from the corresponding cohort. For gene *g*, let 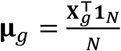denote the vector of mean genotype dosages. The within-gene LD matrix is the SPLR matrix 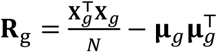 sparse matrix 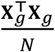 records the mean co-occurrence of rare alleles across individuals, and 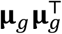 is a rank-1 matrix.

#### Pairwise burden kernel computations

The most computationally intensive step is evaluating pairwise traces between burden and cross-burden kernels. Suppose we have *B* burden weights, represented by the columns of 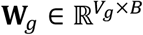. These burden weights generate *B* burden kernels and *B(B−1)/2* cross-burden kernels, for a total of *B(B+1)/2* kernels. Constructing the normal equations therefore requires evaluating tr[**K**_*k*_**RK**_*l*_**R**] for every pair of these kernels, resulting in approximately *B*^*4*^*/8* pairwise calculations. We reduce *O(B*^*4*^) matrix operations to *O(B*^*2*^) by first computing the *B* × *B* burden covariance matrix 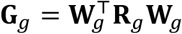 and then computing the trace as follows. Letting **w**_*p,g*_ denote column *p* of **W**_*g*_, there are three cases:

1. Burden kernel 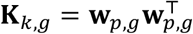. burden kernel 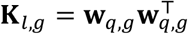.

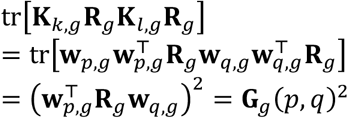
2. Burden kernel 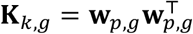vs. cross-burden kernel 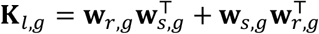

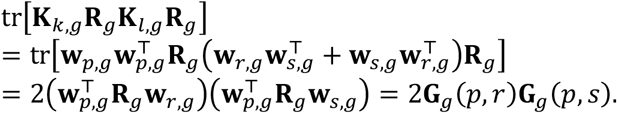
3. Cross-burden kernel 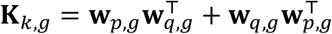vs. cross-burden kernel 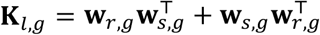

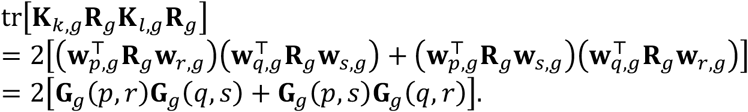

#### Block jackknife computation

Because sufficient statistics are additive across genes under no between-gene LD, each block’s contribution is precomputed once, so the jackknife requires only one exome-wide pass through the LD matrices and annotations.

### Exome-wide association studies and functional annotations

We analyzed rare coding association statistics from UK Biobank and All of Us, using variants with minor allele frequency < 10^−3^. The threshold was selected to minimize tagging the effects of variants outside each gene, following previous work^9^. For UK Biobank, we used Genebass summary statistics^38^; for All of Us, we used All by All v8 Controlled Tier summary statistics^39^. The main joint analysis included 16 UK Biobank continuous traits and 19 All of Us continuous traits in each of the European-like (EUR-like), African-like (AFR-like), and admixed American-like (AMR-like) genetic-similarity groups. For each variant-trait pair, we extracted the marginal effect estimate, its standard error, allele frequency and sample size. Variants were harmonized by GRCh38 coordinate, reference and alternate alleles. We additionally analyzed four UK Biobank binary disease traits (diabetes, asthma, osteoarthritis, and cancer), estimating kernel heritability on the observed scale.

We classified rare coding variants into three broad consequence categories: predicted loss-of-function (pLoF), missense, and synonymous variants, as provided in Genebass^38^ and All by All^39^.

#### Missense variant scores

We analyzed 15 quantitative scores spanning supervised and unsupervised deleteriousness prediction, evolutionary conservation, splicing, mutation rate, and protein language model-based annotations. These included PolyPhen-2^10^, AlphaMissense^14^, ESM-1v^12^, REVEL^88^, PrimateAI-3D^13^, MisFit-S^89^, SIFT^90^, CADD^11,91^, phyloP^92^, and popEVE^16^. Scores were matched by genomic coordinate and reference and alternate alleles. Details about sources and definitions for each score are reported in **Supplementary Table 6**.

#### Primary-sequence substitution annotations

We annotated each missense substitution from its reference and alternate amino acids in the canonical UniProt sequence. Charge-reversing substitutions were Glu↔Lys substitutions, the only single-nucleotide changes between positively and negatively charged amino acids. Hydrophobicity-disrupting substitutions were changes between a hydrophobic (C, L, V, I, M, F, W) and a polar (R, K, E, D, Q, N) amino acid. Large volume-changing substitutions changed side-chain volume by more than 90 Å^3^ (Grantham volumes^93^).

#### Missense structural and functional annotations

We analyzed structural and functional annotations from NetSurfP^94^, PIONEER^21^, and the Genomics 2 Proteins (G2P) portal^95^. These included predicted solvent accessibility, secondary structure and intrinsic disorder, UniProt-annotated transmembrane regions and disulfide bonds^96^, PDB-supported intrachain and interchain contacts, and protein-protein interaction interfaces. Intrinsically disordered residues were defined as those in the highest within-gene quintile of NetSurfP-predicted disorder scores. We mapped residue-level annotations to variants using canonical protein coordinates derived from transcript consequence annotations.

#### Residue burial and protein interaction interface

We retrieved the HINT high-quality binary human interactome^97^ (2024-06 release) and retained protein-protein interactions involving exome-wide significant genes that were supported by at least one PDB co-crystal chain pair. For each PDB chain pair, we retrieved the deposited mmCIF structure from RCSB^98^, mapped PDB residues to UniProt positions using per-residue SIFTS mappings^99^, and computed solvent-accessible surface area using FreeSASA^100^. For each gene, solvent accessibility was computed both for the isolated chain and for the two-chain complex. Residues were annotated as interface residues if they were solvent-exposed in the isolated chain (relative SASA ≥ 15%) and lost at least 1.0 Å^2^ SASA upon complex formation. Residues were annotated as complex-buried if their relative solvent accessibility in the complex was < 25%.

#### Synonymous variant annotations

For synonymous variants, we used five splicing, conservation, deleteriousness, and mutation-rate scores: phyloP^92^, GERP++^101^, CADD^11^, Roulette mutation rate^102^, and SpliceAI^6^ maximum delta score across acceptor gain, acceptor loss, donor gain and donor loss. Details about sources and definitions for each score are reported in **Supplementary Table 6**.

#### Deep mutational scanning (DMS)

We curated four DMS assays with sufficient overlap with rare missense variants in the exome association data in well-powered gene-trait associations: FGFR3-height, GCK-HbA1c, LDLR-LDL cholesterol, and MC4R-BMI. We compiled them from MaveDB (FGFR3^51^, LDLR^53^, 2026-02-06 quarterly Zenodo bulk release)^103^, ProteinGym (GCK^52^, v1.3)^104^, and the Octant MC4R DMS (github.com/octantbio/mc4r-dms^54^). DMS target sequences were aligned to the canonical human UniProt proteome using MMseqs2^105^; protein targets were aligned directly, whereas DNA targets were translated in the three forward reading frames before protein-level alignment. Substitutions were converted to all codon-compatible genomic SNVs, producing (chr:pos:ref:alt) variant IDs for overlap with exome association statistics.

#### Gene-level annotations

Following ref.^9^, we partitioned genes by quintiles of the observed/expected number of unique pLoF variants in gnomAD^47^. The first quintile contains the most constrained genes and the last quintile the least. We estimated heritability enrichment across these quintiles separately for pLoF variants, all missense variants, and AlphaMissense-prioritized missense variants (the highest-scoring missense variants within each gene, retaining up to the number of pLoF variants in that gene).

### Functionally informed kernel heritability

We fit KASM models separately to pLoF, missense, and synonymous variants, using random-effects estimators for subthreshold genes, and fixed-effects estimators for significant genes. MAF-stratified models used bins of 0-10^−5^, 10^−5^-10^−4^, 10^−4^-10^−3^, with matching diagonal overdispersion kernels for the same bins. All models used five gnomAD pLoF observed/expected bins for gene-level scaling. Unless noted, pLoF and synonymous variants used one constant weight per MAF bin.

#### Naïve

To estimate heritability under a conventional burden assumption, a single constant burden weight was assigned separately for pLoF and missense variants. The random-effects estimator modeled the corresponding rank-one burden kernel across subthreshold genes, and the fixed-effects estimator fitted the same one-dimensional burden score separately in each significant gene.

#### BHR-like

To approximate BHR’s functional and MAF stratification, missense variants were divided by the three MAF bins and binary PolyPhen-2 classification, yielding separate burden weights for damaging and benign variants within each MAF bin.

#### KASM Baseline

To model quantitative differences in missense variant deleteriousness, we used four burden weights: a constant weight, PolyPhen-2, ESM-1v, and AlphaMissense. We stratified each weight by the three MAF bins, yielding 12 burden weights.

KASM Baseline + Annotation

For a binary annotation, we added one annotation-specific weight per MAF bin. The subthreshold model included all burden and pairwise cross-burden kernels from the expanded weight set, and the significant-gene model included the same weights in the fixed-effects burden matrix.

KASM Baseline x Exon

Variants were assigned to coding exons using GENCODE v39 coding-sequence annotations^106^, and each exon was treated as an aggregation unit. Baseline weights were restricted to variants in each exon, and every exon inherited its parent gene’s constraint quintile. The random-effects model was fit across exon-level units in subthreshold genes. In significant genes, fixed-effects heritability was estimated for each exon and summed within each gene; gene significance was still defined by the gene-level burden test.

#### KASM Baseline + Indep

To estimate heritability not represented by the gene- or exon-level burden kernels, we implemented estimators that allowed a more flexible variant effect distribution. For subthreshold genes, we added an identity kernel for each of pLoF, missense and synonymous variant classes to model independent variant-effect variance not represented by the burden and cross-burden kernels. Because this kernel is partially collinear with MAF-stratified overdispersion kernels, we constrained the overdispersion coefficients to be shared across the pLoF, missense, and synonymous variants when analyzing each trait. This was supported by similar coefficients across classes (**Supplementary Figure 8**). To further improve identifiability, we first fit KASM Baseline, fixed its gene-scaling coefficients, and then jointly estimated the burden-kernel, identity-kernel, and overdispersion coefficients. This procedure encodes the assumption that constrained genes are equally enriched for variant effects which are or are not aligned with functional annotations. For significant genes, the baseline burden model was replaced by the variant-level fixed-effects estimator.

#### KASM Baseline + Indep. + Position

To estimate effect concordance among proximal variants, we extended KASM Baseline + Indep. by adding one binary position kernel at a time (see above).

### Simulations

We simulated rare coding effects as the sum of gene-level burden, exon-level missense burden, independent missense and overdispersion effects, and generated the associational effect vector as 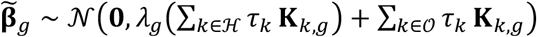. Conditional on 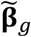 and within-gene LD matrix **R**_*g*_, we simulated marginal variant association effects according to 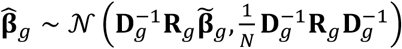.

Heritable kernel coefficients, gene-scaling coefficients, and overdispersion coefficients were obtained by fitting “KASM Baseline + Indep.” to height summary statistics from EUR-like All of Us participants. Variant annotations, allele frequencies, sample sizes, and within-gene LD matrices were from All of Us EUR-like participants. To introduce model misspecification, we simulated gene-level sparsity that deviates from the KASM model; we randomly selected 1% of genes and multiplied their effect variance by 25, 50, or 100, then rescaled all causal effects to keep total heritability fixed.

Each simulation setting used 100 replicates. We fit BHR-like, KASM Baseline, KASM Baseline x Exon and KASM Baseline + Indep. models to estimate their corresponding heritability estimands. For enrichment, we evaluated AlphaMissense-ranked sets within each gene: the most deleterious variants, variants centered on the 10th percentile, the most benign variants, and a random set of the same size.

We also performed individual-level null simulations with common-variant effects but no rare coding effects in EUR-like, AFR-like, and AMR-like All of Us cohorts, as described in ref.^39^. The common-variant genetic component was sampled from a genetic relationship matrix and scaled to 50% heritability; independent environmental noise produced standardized continuous phenotypes. We calculated rare-variant association statistics from each phenotype and fit BHR-like, KASM Baseline, KASM Baseline x Exon and KASM Baseline + Indep. Five replicates were analyzed for each genetic-similarity group.

### Benchmarking missense deleteriousness predictors

For each of the 15 missense deleteriousness predictors, we ranked variants within each gene and defined the top decile as the variants predicted to be most deleterious. Variants in the top decile were analyzed using a binary annotation with KASM Baseline + Annotation, with one annotation-specific weight per MAF bin.

We constructed a KASM composite score from the fitted KASM Baseline covariance. For variant *v* in gene *g*, the score was the diagonal entry *K*_*g,vv*_, representing fitted per-allele effect variance. Coefficients were estimated in meta-analysis across traits. To prevent overfitting, separate models were trained on even- and odd-chromosome genes, and each model scored variants on the opposite chromosome set. We excluded each trait’s exome-wide significant genes in model fitting. Variants were ranked within genes, and the top decile was defined and evaluated with the same enrichment procedure as individual predictors. We also constructed an alternative composite score with 2*f*_*v*_(1 − *f*_*v*_)*K*_*g,vv*_, representing predicted per-variant heritability. For this analysis, we defined heritability enrichment as the fraction of missense heritability attributable to the selected variants divided by their fraction of missense variants, rather than their fraction of missense heterozygosity.

For the DMS benchmark, each gene-trait comparison was restricted to variants observed in the association data with complete DMS, PolyPhen-2, ESM-1v, and AlphaMissense scores. Within the shared variant set, each score was divided into quartiles, and quartile enrichment was estimated with the fixed-effects estimator.

### Structural and functional enrichment analyses

Each binary structural or functional annotation was tested separately with KASM Baseline + Annotation. Enrichment was estimated independently in significant and subthreshold genes and then meta-analyzed across traits. We tested whether enrichment differed from one with two-sided Wald tests, Bonferroni-corrected across the 20 annotations.

### Context-dependent missense variant effects

We performed context-dependent enrichment analyses for missense variants in exome-wide significant gene-trait associations using the fixed-effects KASM estimator. For each pair of a structural or functional context and a substitution class, we defined two binary annotations within the context: variants with (Yes) and without (No) the substitution class. Each annotation was analyzed separately as a binary annotation on top of the KASM Baseline model, and enrichment was meta-analyzed across traits.

### Effect concordance of variant pairs

To evaluate whether nearby coding variants have more concordant phenotypic effects, we constructed binary position kernels representing proximity in the primary sequence or three-dimensional protein structure. One-dimensional distance was defined as the absolute difference in coding-sequence position. For three-dimensional proximity, we used AlphaFold-predicted structures and GRCh38-to-UniProt residue mappings released by Nason et al.^58^. We calculated the minimum distance between side-chain heavy atoms of each residue pair, excluding glycine and residues with incomplete side chains. For proteins represented by multiple overlapping AlphaFold segments, we used the minimum distance across all segments containing both residues. Each binary relationship was fitted separately as a position kernel in the KASM Baseline+ Indep. + Position model, and effect concordance was estimated from the resulting covariance as described above. We performed several secondary analyses. We included an independent-effect kernel restricted to variants participating in the position kernel. We defined separate position kernels for pairs of residues within the same α-helix or β-strand at each residue offset from 0 to 6. For AlphaMissense-stratified concordance, variants with scores above each threshold (0.564, 0.8, or 0.9) were classified as high, and all other scored variants as other. We fitted separate position kernels for high-high, high-other, and other-other variant pairs.

### Characterizing functional allelic series

To characterize functionally defined allelic series within individual genes, we applied the fixed-effects estimator separately to each significant gene-trait association. We fit KASM Baseline + Annotation and tested enrichment greater than one with a one-sided Wald test.

For the *PIEZO1*-HbA1c example, we used UK Biobank exome association statistics. The *PIEZO1* trimer structure was obtained from the cryo-EM structure PDB 8YEZ. We queried PDBe PISA for assembly interface 1 and extracted hydrogen bonds connecting subunits. To estimate heritability enrichment, we built a binary annotation for missense variants at PISA-defined interchain hydrogen-bond residues and fit KASM Baseline + Annotation.

For the *FGFR3*-height example, we used UK Biobank exome association statistics. We mapped FGFR3 missense variants to canonical protein coordinates and defined the transmembrane helix as UniProt residues 376-396 and the kinase domain as UniProt residues 472-761. For the kinase domain, we used the mapped FGFR3 DMS activation assay restricted to kinase-domain variants overlapping the exome association data and assigned these variants to DMS quartiles. To estimate the heritability enrichment for the transmembrane helix and the kinase-domain DMS quartiles, we partitioned FGFR3 missense variants into six non-overlapping subsets: transmembrane-helix variants, four DMS-scored kinase-domain quartiles, and all remaining missense variants. We then fit a single KASM Baseline fixed-effects model in which each Baseline burden weight was fitted separately within each subset and decomposed the gene’s heritability additively across subsets. The total heritability from this model and the total heterozygosity of all FGFR3 missense variants were used as denominators when estimating heritability enrichment for each subset.

## Data Availability

UK Biobank rare-variant association summary statistics are available from Genebass (https://genebass.org). All of Us All by All v8 Controlled Tier summary statistics are available to authorized researchers through the All of Us Researcher Workbench (https://workbench.researchallofus.org). All data produced in the present study are available upon reasonable request to the authors.

## Data availability

UK Biobank rare-variant association summary statistics are available from Genebass (https://genebass.org). All of Us All by All v8 Controlled Tier summary statistics are available to authorized researchers through the All of Us Researcher Workbench (https://workbench.researchallofus.org).

## Code availability

KASM source code is available at github.com/kangchenghou/kasm-pkg.

## Acknowledgements

We thank Alkes Price for helpful discussions. This research has been conducted using the UK Biobank Resource under Application Number 86805. We thank UK Biobank and All of Us participants for their contributions, without whom this research would not have been possible. We also thank the National Institutes of Health’s All of Us Research Program for making available the participant data examined in this study. This research was supported by NIH NIGMS R35 GM155278 (L.J.O.).

## Supplementary Figures

**Supplementary Figure 1.**
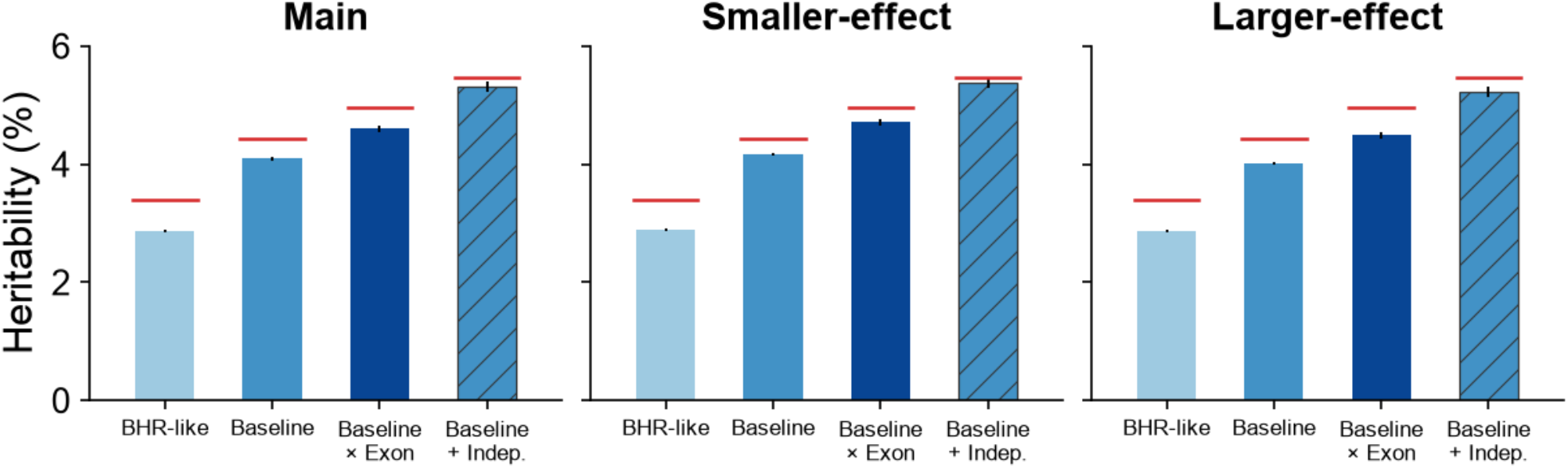
Heritability estimates across simulation settings. Heritability estimates under the BHR-like, KASM Baseline, KASM Baseline x Exon and KASM Baseline + Indep. models. Gene-level effect variances are scaled by 50 (Main), 25 (Smaller-effect) or 100 (Larger-effect) in a random 1% of genes and by 1 in all remaining genes. Bars show means across 100 replicates, and red line segments show simulated estimands. Error bars show standard errors.

**Supplementary Figure 2.**
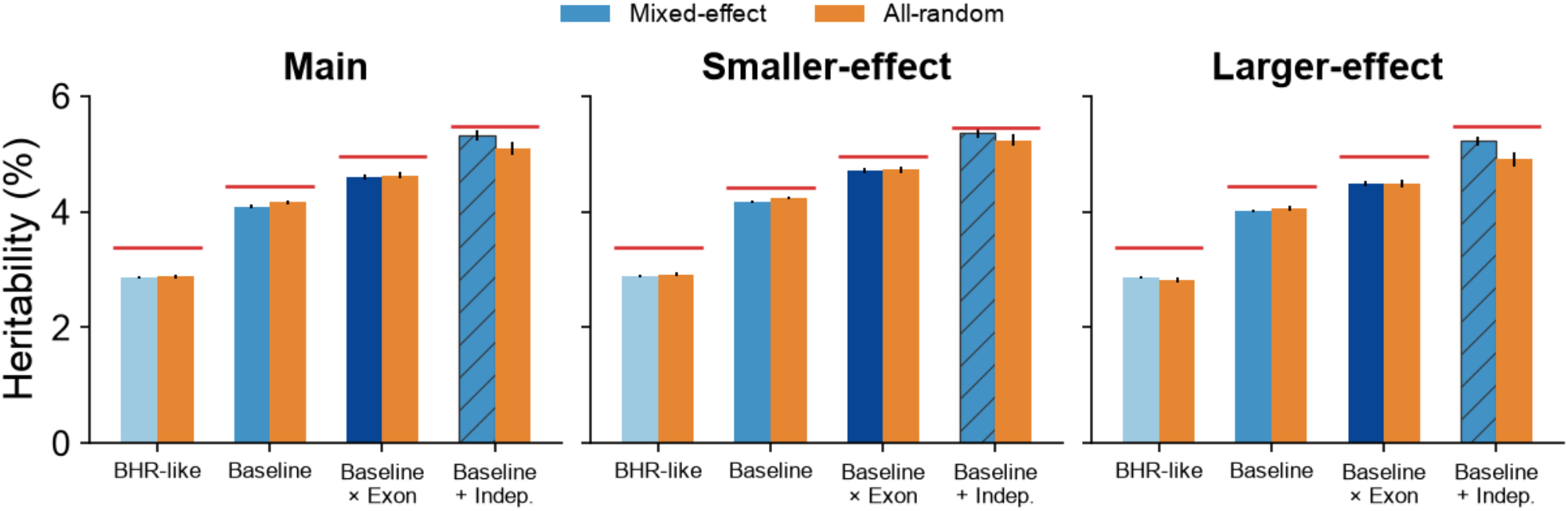
Mixed-effects versus all-gene random-effects estimation. Gene-level effect variances are scaled by 50 (Main), 25 (Smaller-effect) or 100 (Larger-effect) in a random 1% of genes and by 1 in all remaining genes. The default mixed-effects estimator (Mixed-effect) combines fixed effects for exome-wide significant gene-trait associations with random effects for subthreshold genes; the alternative (All-random) treats all genes as random effects. Bars show means across 100 replicates, and red line segments show simulated estimands. Error bars show standard errors.

**Supplementary Figure 3.**
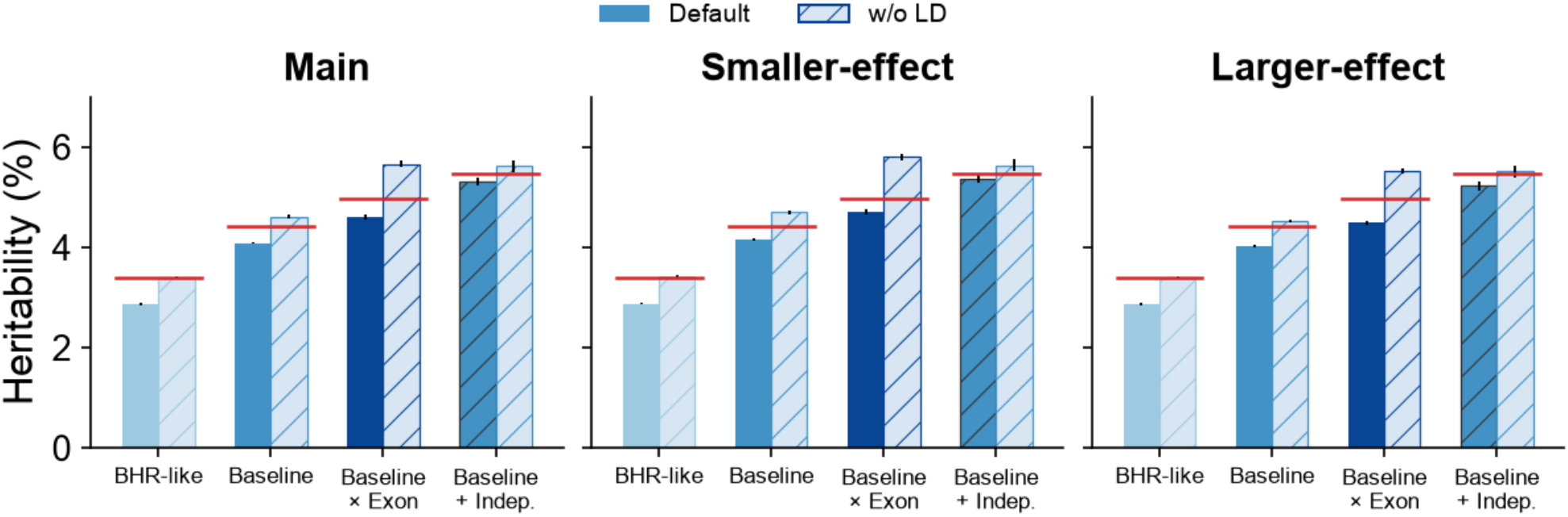
Effect of accounting for within-gene LD on heritability estimates. Gene-level effect variances are scaled by 50 (Main), 25 (Smaller-effect) or 100 (Larger-effect) in a random 1% of genes and by 1 in all remaining genes. Summary statistics were simulated using empirical within-gene rare-variant linkage disequilibrium (LD) from All of Us. Default fits use the corresponding LD matrices; ‘w/o LD’ fits treat variants as independent. Bars show means across 100 replicates, and red line segments show simulated estimands. Error bars show standard errors.

**Supplementary Figure 4.**
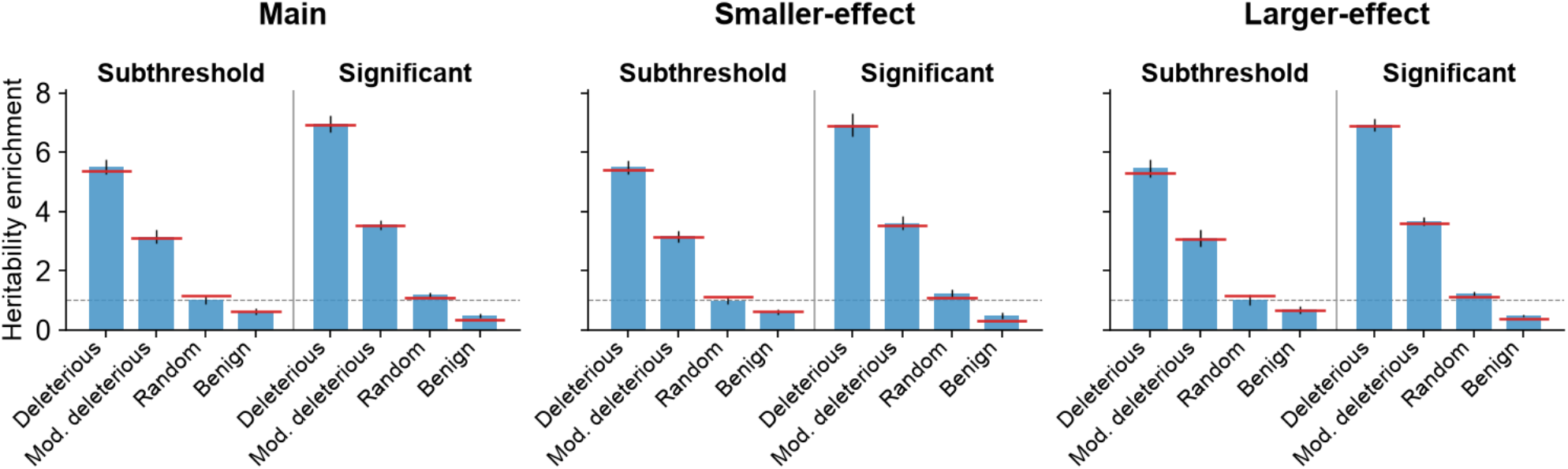
Heritability enrichment estimates across simulation settings. Gene-level effect variances are scaled by 50 (Main), 25 (Smaller-effect) or 100 (Larger-effect) in a random 1% of genes and by 1 in all remaining genes. Annotation sets contain 2% of missense variants within each gene: the most deleterious, those centered on the 10th percentile, a random sample or the most benign. Results are shown separately for subthreshold genes and exome-wide significant gene-trait associations. Bars show enrichment pooled across 100 simulation replicates, and red segments show simulated enrichment. Error bars show standard errors.

**Supplementary Figure 5.**
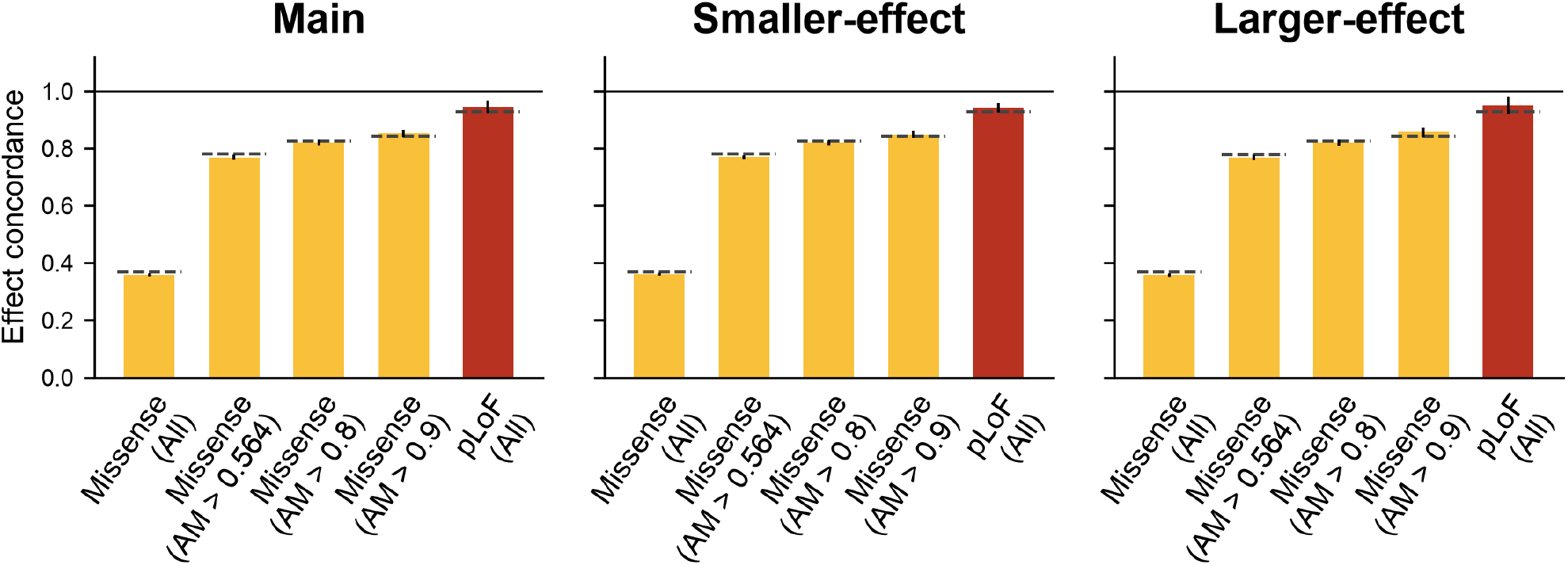
Accuracy of effect concordance estimation in simulations. Gene-level effect variances are scaled by 50 (Main), 25 (Smaller-effect) or 100 (Larger-effect) in a random 1% of genes and by 1 in all remaining genes. Concordance is evaluated for all within-gene missense pairs, missense pairs with both AlphaMissense scores above 0.564, 0.80 or 0.90, and all within-gene pLoF pairs. Bars show concordance pooled across 100 replicates, and dashed segments show true causal-effect concordance. Error bars show standard errors.

**Supplementary Figure 6.**
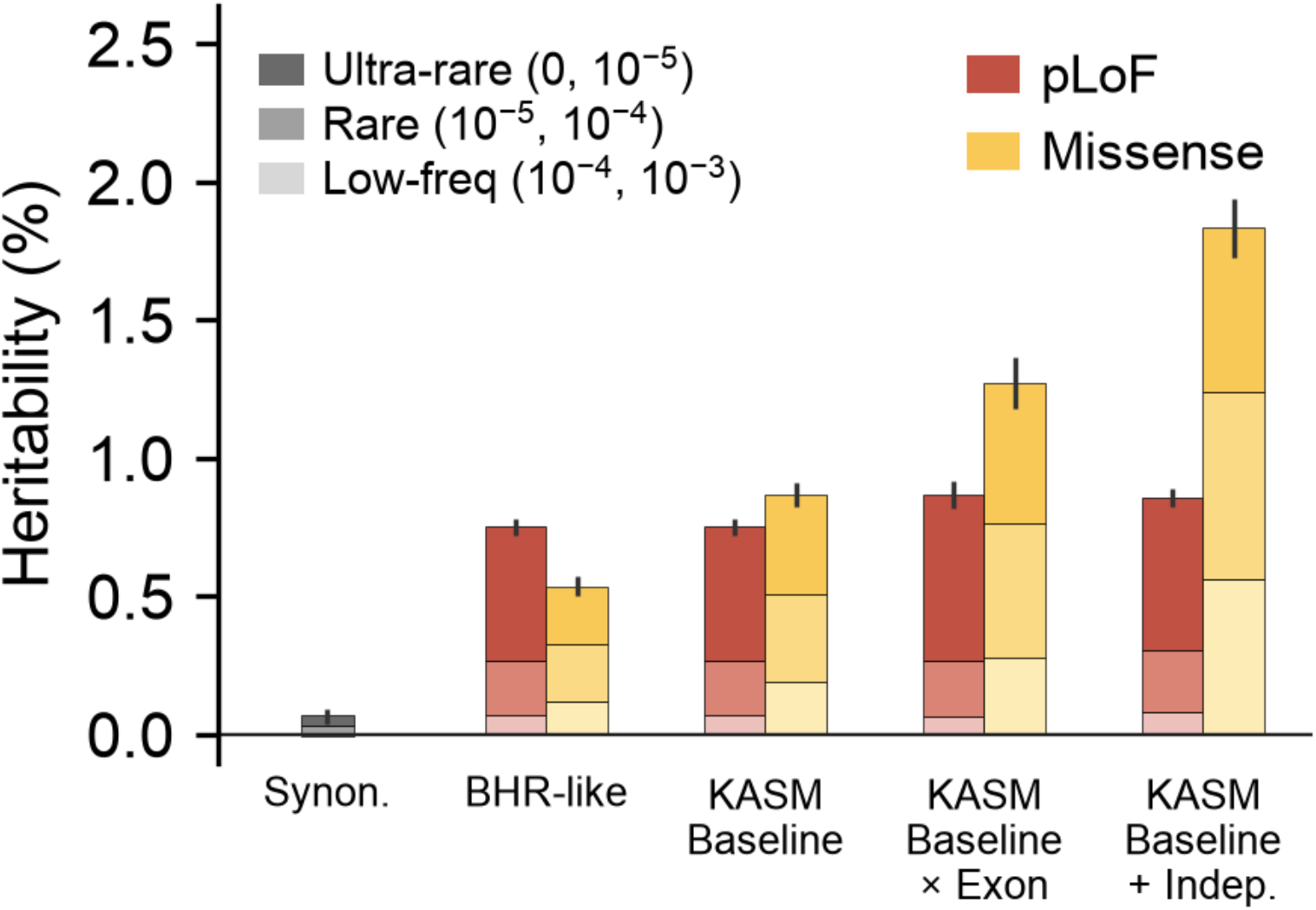
Rare coding heritability across the allele-frequency range. Heritability estimates from **Figure 2b** for synonymous, pLoF and missense variants, decomposed into ultra-rare, rare, and low-frequency bins. Estimates are meta-analyzed across UK Biobank and All of Us traits. Error bars show standard errors of heritability summed across MAF bins.

**Supplementary Figure 7.**
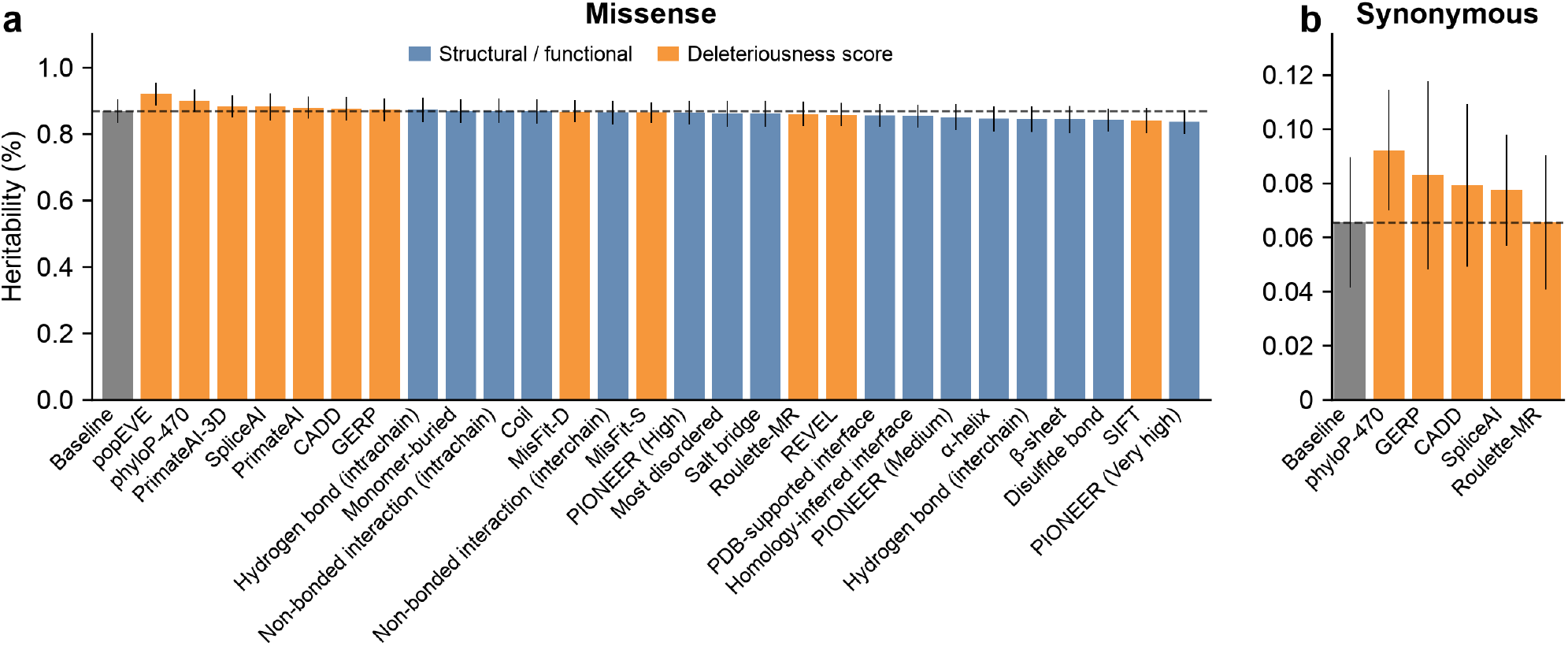
Heritability estimates after including additional annotations. Each bar shows total mixed-effects heritability after adding the indicated annotation to KASM Baseline, meta-analyzed across UK Biobank and All of Us traits for missense variants **(a)** and synonymous variants **(b)**. Adding SpliceAI did not increase synonymous heritability; variants with high SpliceAI scores were sparse, with only 0.47% and 0.17% of synonymous variants having scores ≥0.5 and ≥0.8, respectively. The gray bar and horizontal dashed line show KASM Baseline without an additional annotation. Error bars show standard errors.

**Supplementary Figure 8.**
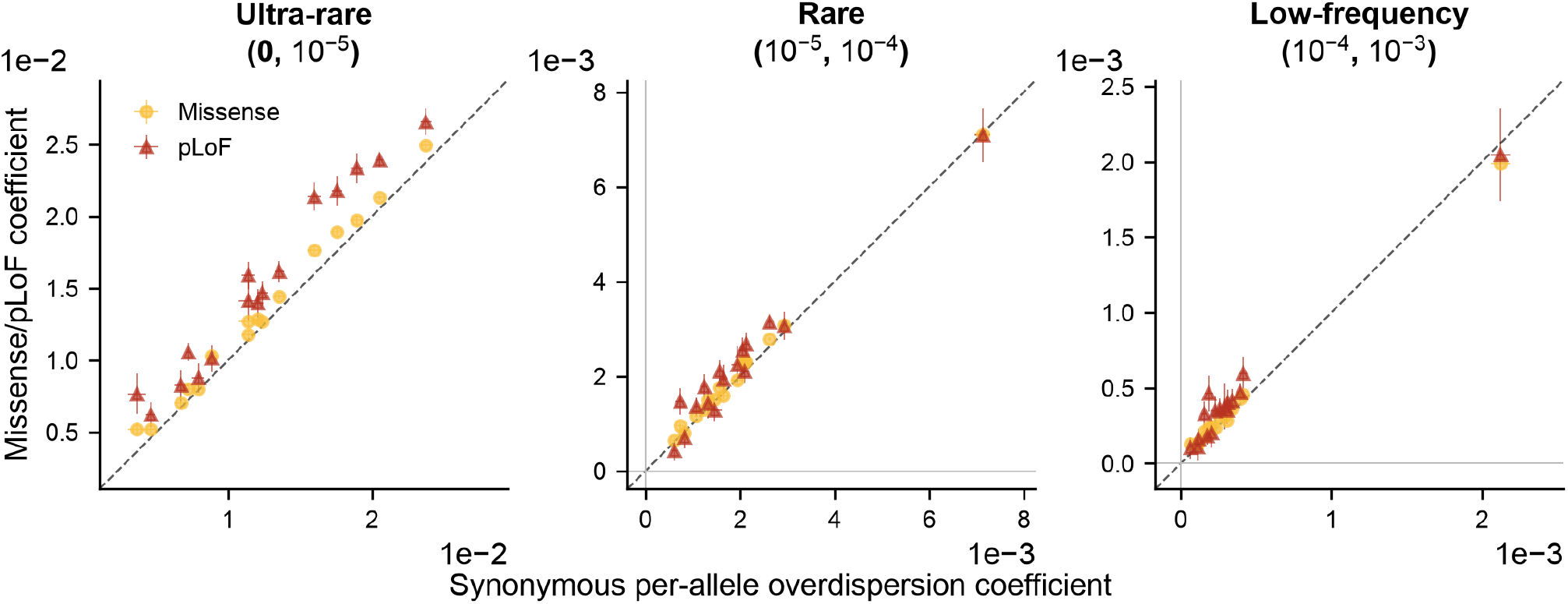
Overdispersion effect estimates across allele-frequency and variant classes. Per-trait per-allele overdispersion coefficients for missense and pLoF variants are compared with synonymous coefficients across 16 UK Biobank traits and three MAF bins (columns). Each point pairs the synonymous coefficient (x axis) with the corresponding missense or pLoF coefficient (y axis) for the same trait and MAF bin. Dashed diagonals denote y = x. Similar coefficients across variant classes are consistent with these terms primarily tagging residual population stratification rather than biological overdispersion. Error bars show standard errors.

**Supplementary Figure 9.**
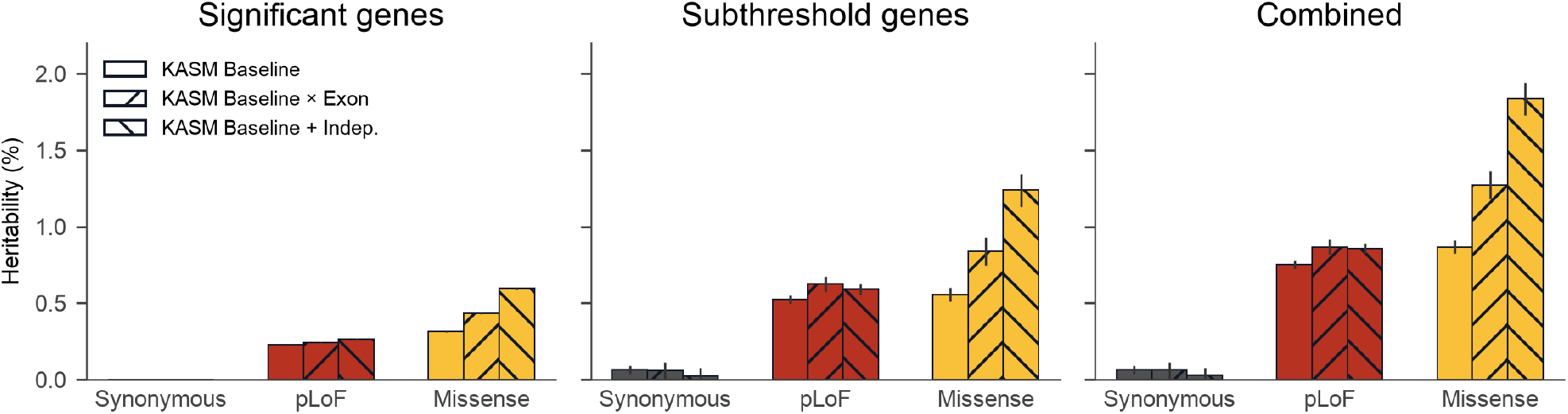
Consistent increases in heritability estimates with more flexible models in significant and subthreshold genes. Panels show heritability estimates for exome-wide significant gene-trait associations, subthreshold genes and their combined mixed-effects totals, meta-analyzed across UK Biobank and All of Us traits. Error bars show standard errors.

**Supplementary Figure 10.**
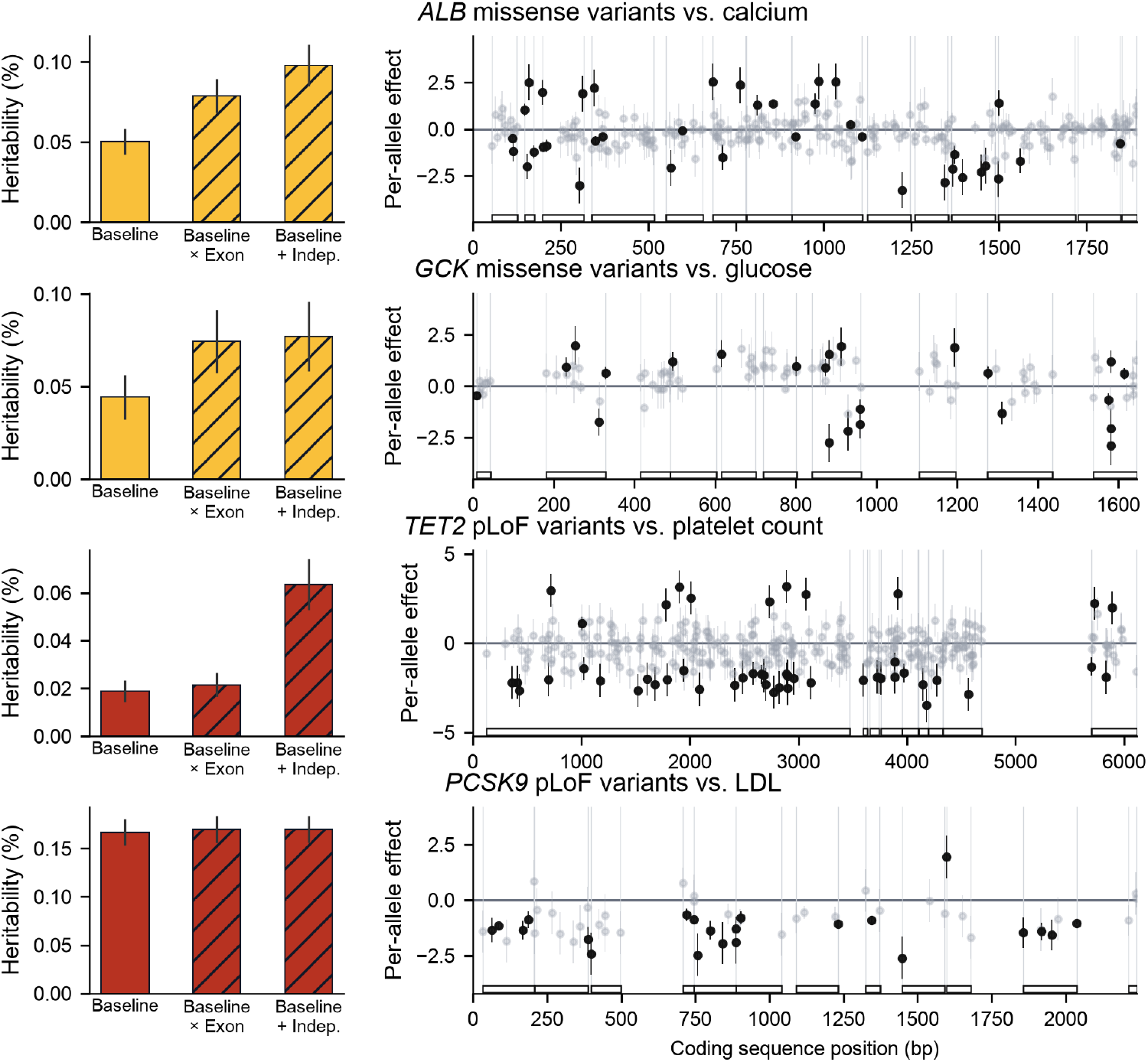
Selected gene-trait associations. Rows show associations for *ALB* missense variants with calcium, *GCK* missense variants with glucose, *TET2* pLoF variants with platelet count and *PCSK9* pLoF variants with LDL. Left, kernel heritability estimated using KASM Baseline, KASM Baseline x Exon and KASM Baseline + Indep.; error bars show standard errors. Right, per-allele effect estimates with standard errors ordered by coding-sequence position; black points denote nominal associations (*P* < 0.05), gray points denote the remaining variants and outlined tracks show exon positions. *ALB* (encoding Albumin) missense variants and serum calcium. Albumin is the main protein-bound carrier of serum calcium; total calcium depends strongly on albumin concentration. *ALB* missense variants could therefore affect total calcium through distinct mechanisms, by changing albumin abundance or stability, or by altering calcium binding^107^. *TET2* pLoF variants and platelet count. Variant-level effects are heterogeneous in direction and are not well represented by a shared gene- or exon-level effect. *TET2* is a common driver of clonal hematopoiesis^46^, and somatic *TET2* mutations may be detected in blood-derived sequencing data. A previous analysis found that the rare-variant association between *TET2* and platelet count was largely attenuated after accounting for variants annotated as somatic^45^, suggesting that somatic clonal hematopoiesis may contribute to the heterogeneous associations observed here.

**Supplementary Figure 11.**
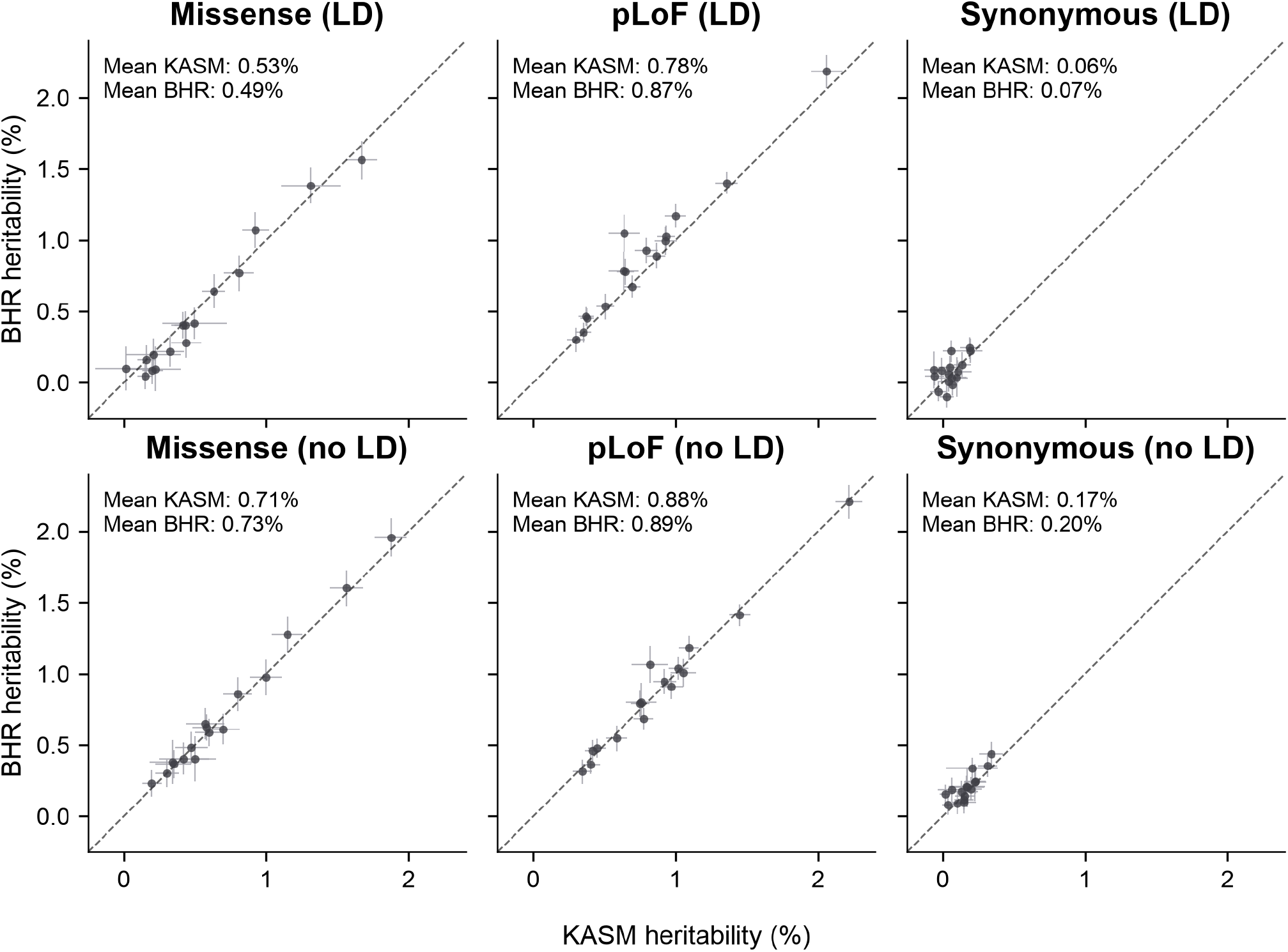
Comparison of KASM and BHR heritability estimates. Mixed-effects heritability estimates from KASM and BHR are compared across 16 UK Biobank traits for missense, pLoF and synonymous variants (columns). Both methods use the identical summary statistics inputs and matched variant categories: pLoF, synonymous, PolyPhen-2-damaging missense and PolyPhen-2-benign missense variants, each divided into three MAF bins. In the top row (LD), KASM models empirical within-gene LD directly, whereas BHR applies an aggregate LD correction; in the bottom row (no LD), KASM uses identity LD and the BHR LD correction is disabled. Each point represents one trait, panel annotations report mean estimates across traits, and dashed diagonals denote y = x. Error bars show standard errors.

**Supplementary Figure 12.**
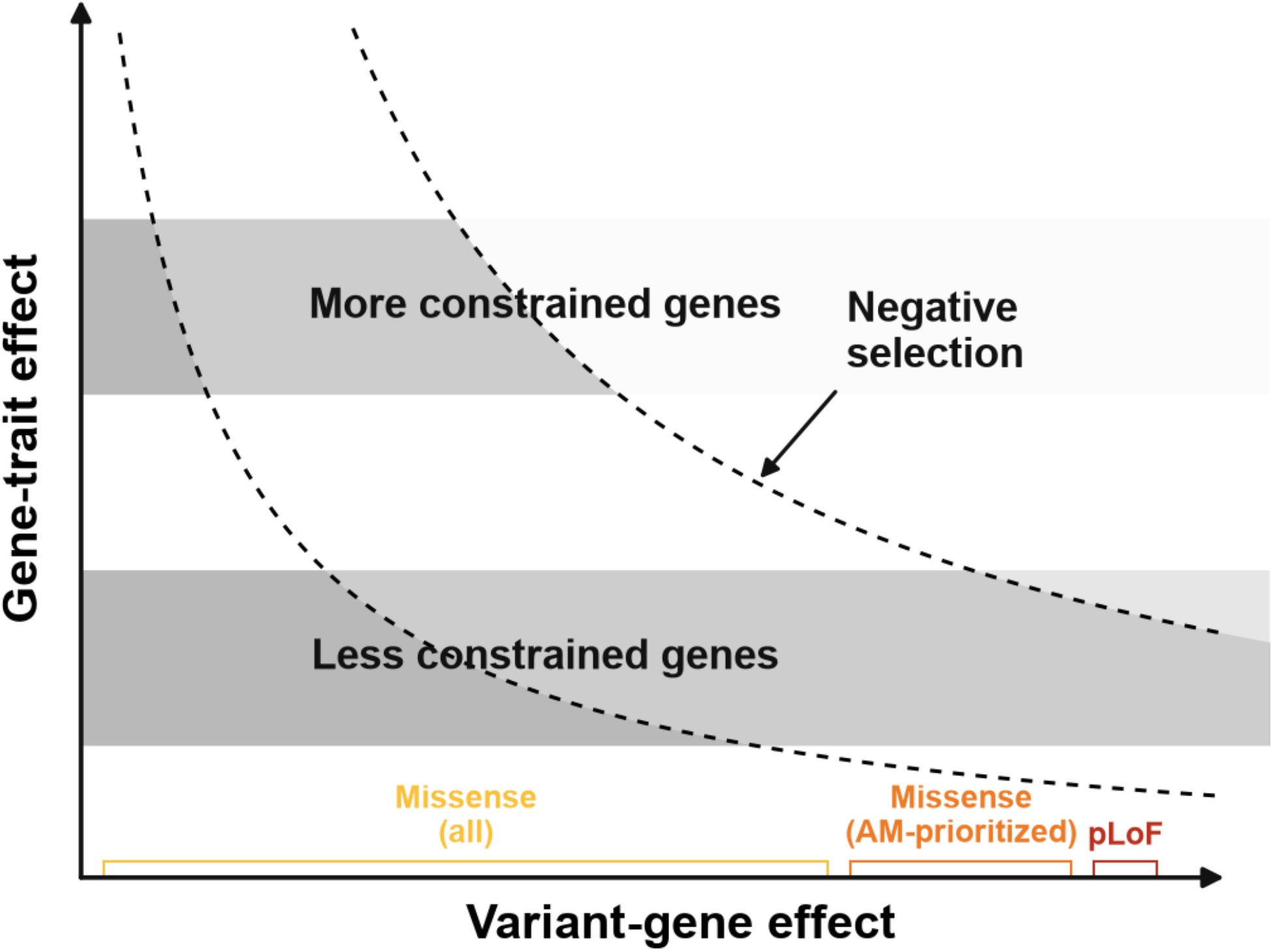
Selection-induced flattening for missense variants. The x-axis represents the variant→gene effect of a coding variant and the y-axis represents the gene→trait effect of the affected gene. Dashed curves show variants with the same product of variant→gene x gene→trait effects, corresponding to similar observed trait effects. Under negative selection, variants with large variant→trait effects are depleted; the gray shading corresponds to the resulting variant density. When all missense variants are used to examine heritability enrichment across gene-constraint bins, missense variants in more constrained genes are increasingly drawn from variants with smaller variant→gene effects, therefore with smaller variant→trait effects and flattening the relationship with gene constraint. Restricting to AlphaMissense-prioritized missense variants, or using pLoF variants, conditions on the variant→gene effects, and the corresponding heritability enrichment results better reflect the gene→trait effect.

**Supplementary Figure 13.**
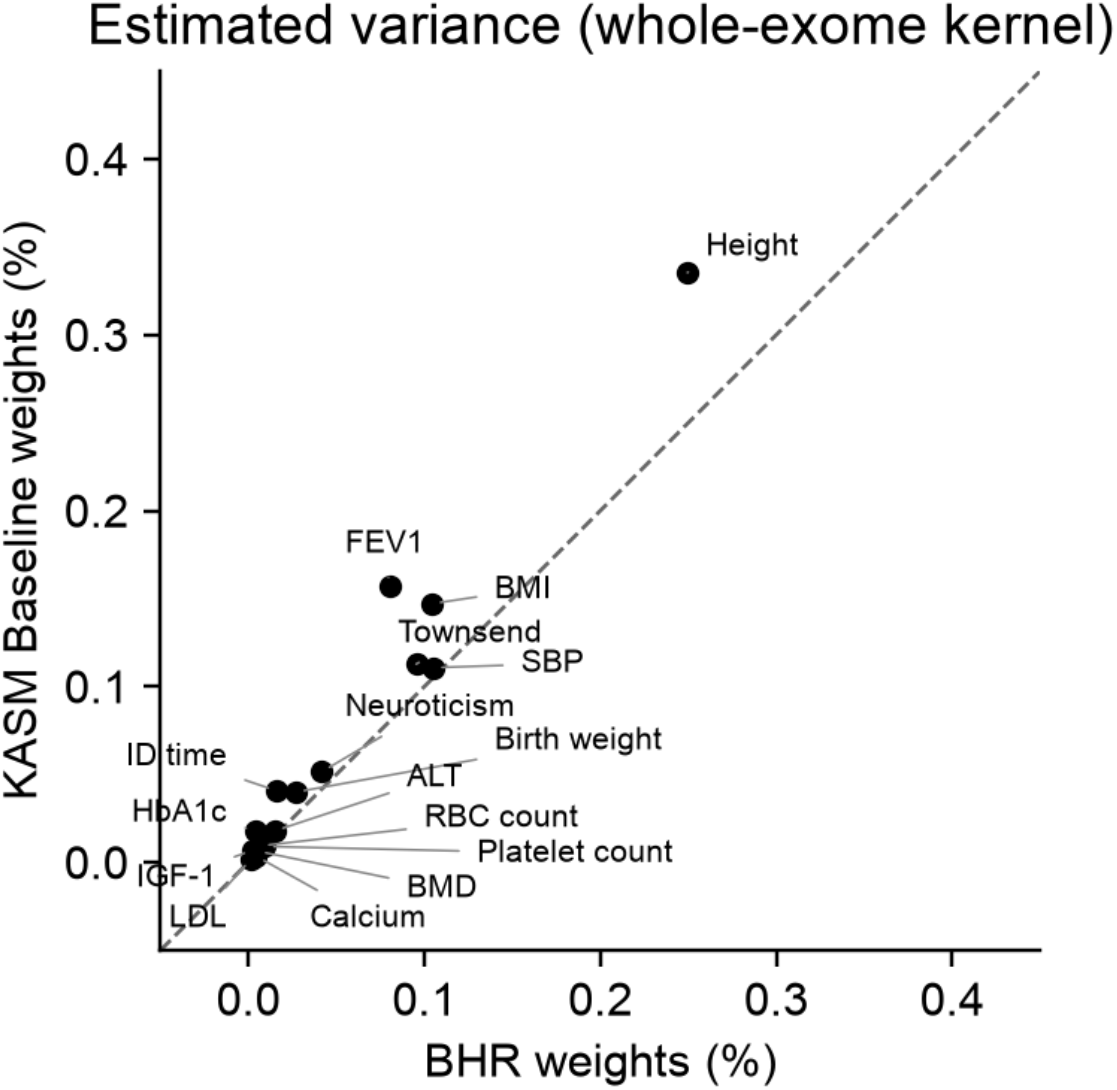
Assessment of residual population stratification aligned with functional annotations. Exome-wide diagnostic estimates obtained by pooling variants across genes and fitting KASM with either BHR or KASM burden weights (Methods). This analysis assesses effect patterns shared across genes that could reflect residual population stratification, although biological effects may also contribute. Estimates are expressed as percentages of phenotypic variance.

**Supplementary Figure 14.**
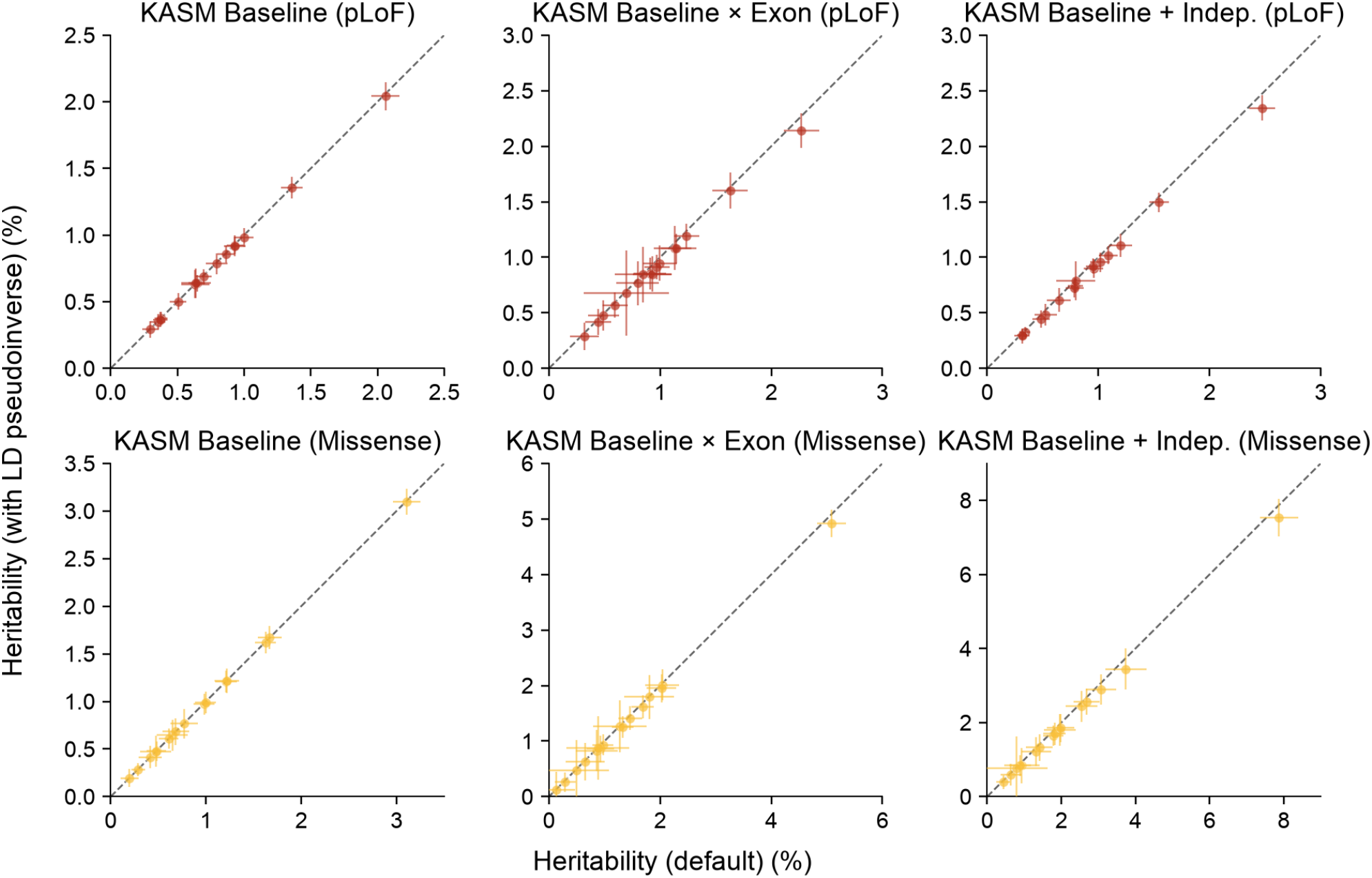
Sensitivity of kernel heritability estimates to inclusion of the genotype covariance pseudoinverse. Comparison of kernel heritability estimates with and without the genotype covariance pseudoinverse added as an overdispersion kernel for KASM Baseline, KASM Baseline x Exon, and KASM Baseline + Indep. Each point represents a UK Biobank trait-variant-class combination for missense or pLoF variants. Gene coefficients were held fixed at their original values, and fixed-effects estimates were unchanged. Horizontal and vertical error bars indicate standard errors for the original and augmented models, respectively (**Methods**).

**Supplementary Figure 15.**
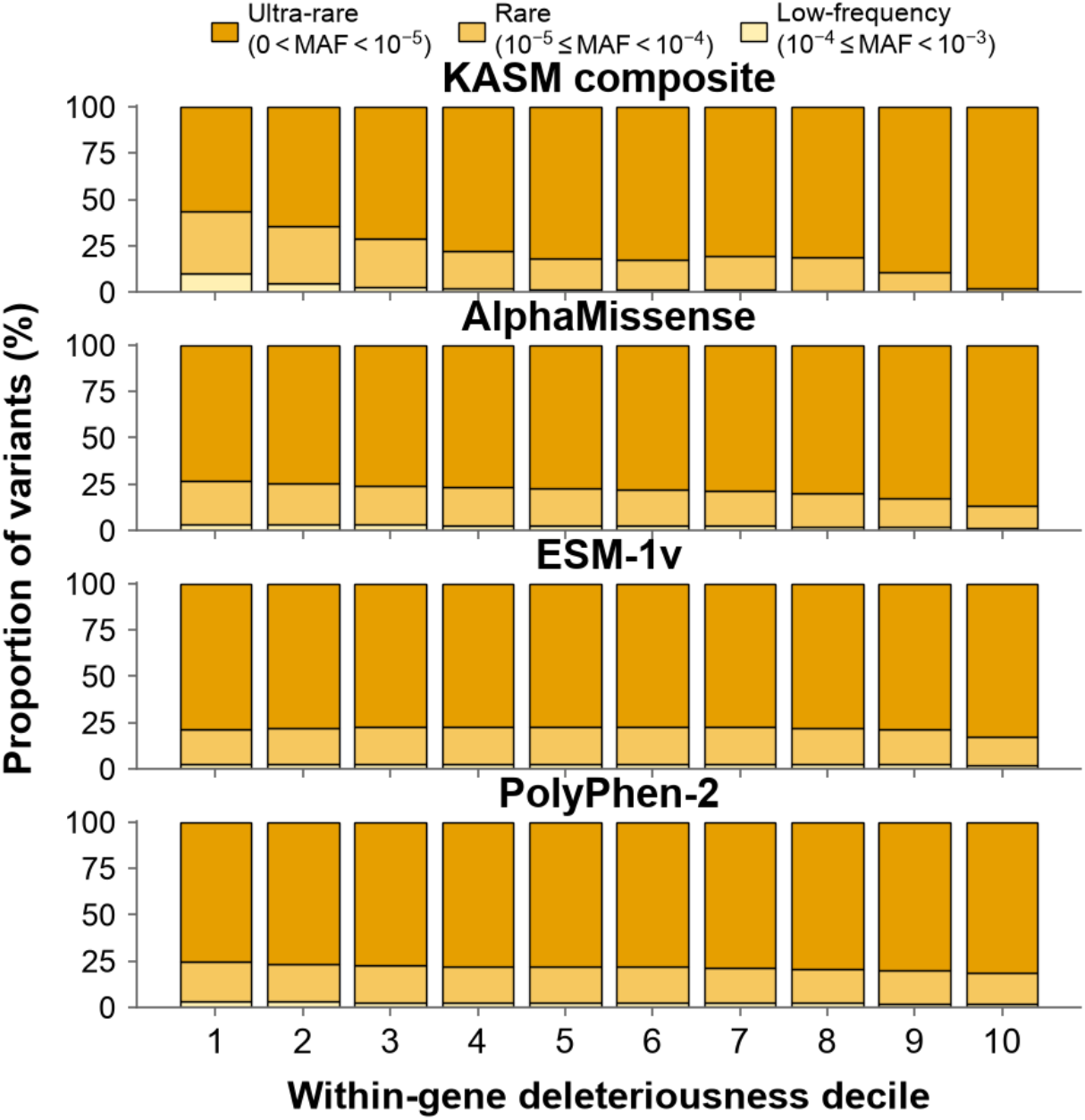
Allele-frequency composition across deleteriousness scores. Missense variants were ranked within gene using KASM composite, AlphaMissense, ESM-1v or PolyPhen-2 and divided into deleteriousness deciles, from least deleterious (1) to most deleterious (10). Within each panel, stacked bars show the proportion of variants in the three MAF bins.

**Supplementary Figure 16.**
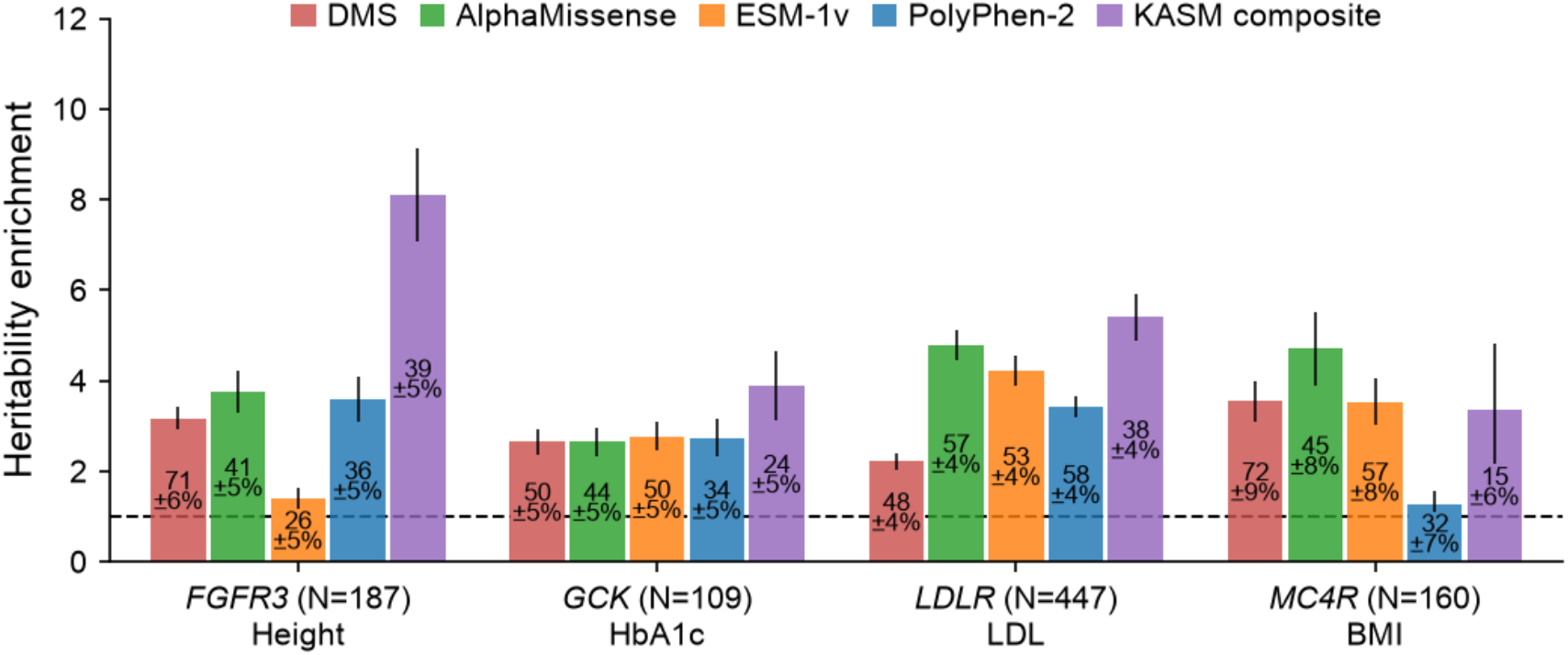
Heritability enrichment of variants prioritized by deep mutational scanning and computational predictors. Top-quartile enrichment is shown for DMS, AlphaMissense, ESM-1v, PolyPhen-2 and the KASM composite score across *FGFR3*-height, *LDLR*-LDL cholesterol, *GCK*-HbA1c, and *MC4R*-body mass index. Bars show heritability enrichment relative to the corresponding heterozygosity share, and error bars show standard errors. Labels within bars report the percentage of gene heritability attributed to the top quartile ± standard error. N denotes the number of variants shared across predictors.

**Supplementary Figure 17.**
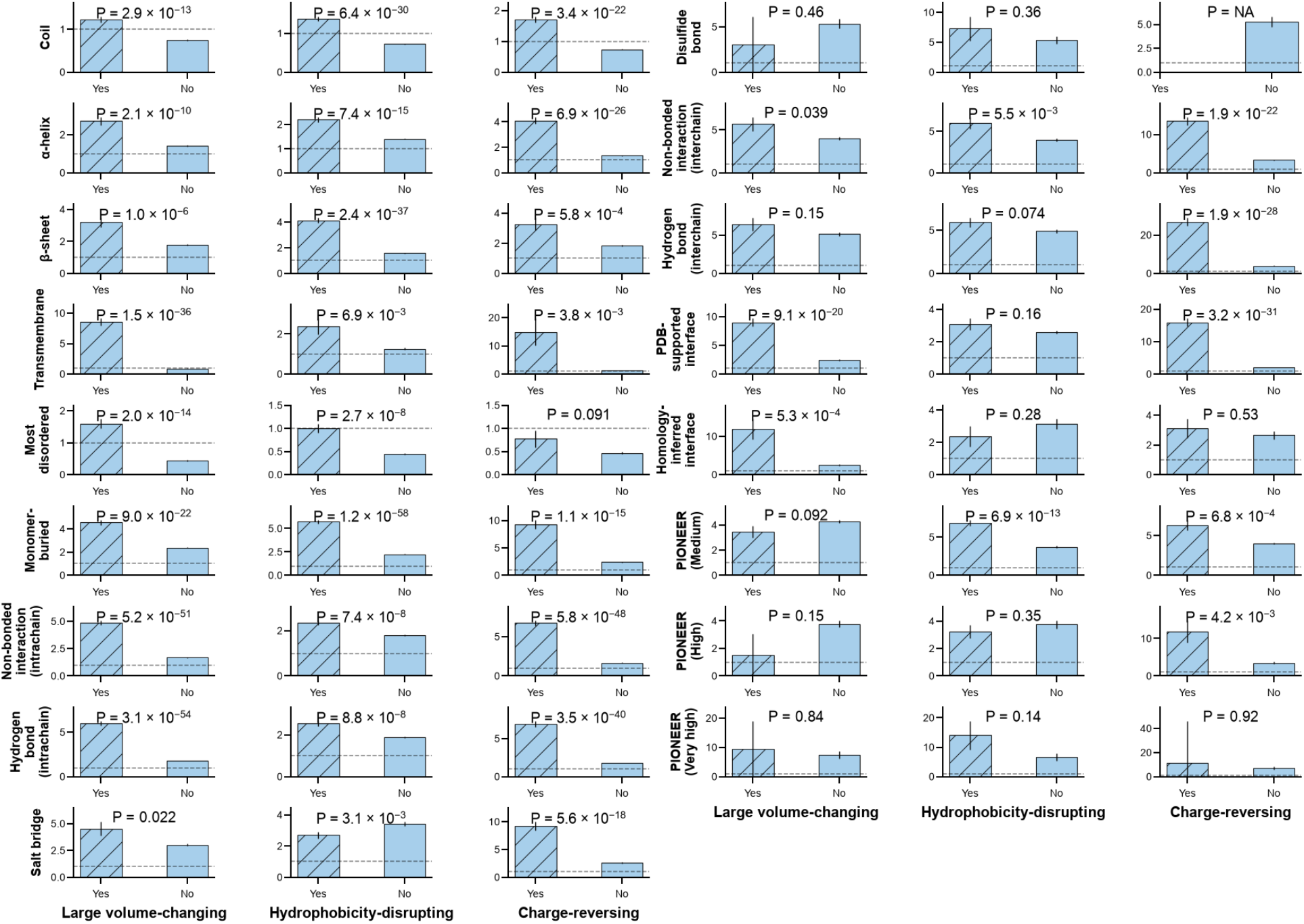
Context-dependent structural and functional enrichment across amino-acid substitution classes. Heritability enrichment is shown for large volume-changing, hydrophobicity-disrupting and charge-reversing substitutions among exome-wide significant gene-trait associations, stratified by the selected protein structural or functional annotation. Two-sided *P* values compare enrichment within and outside each substitution class. Error bars show standard errors.

**Supplementary Figure 18.**
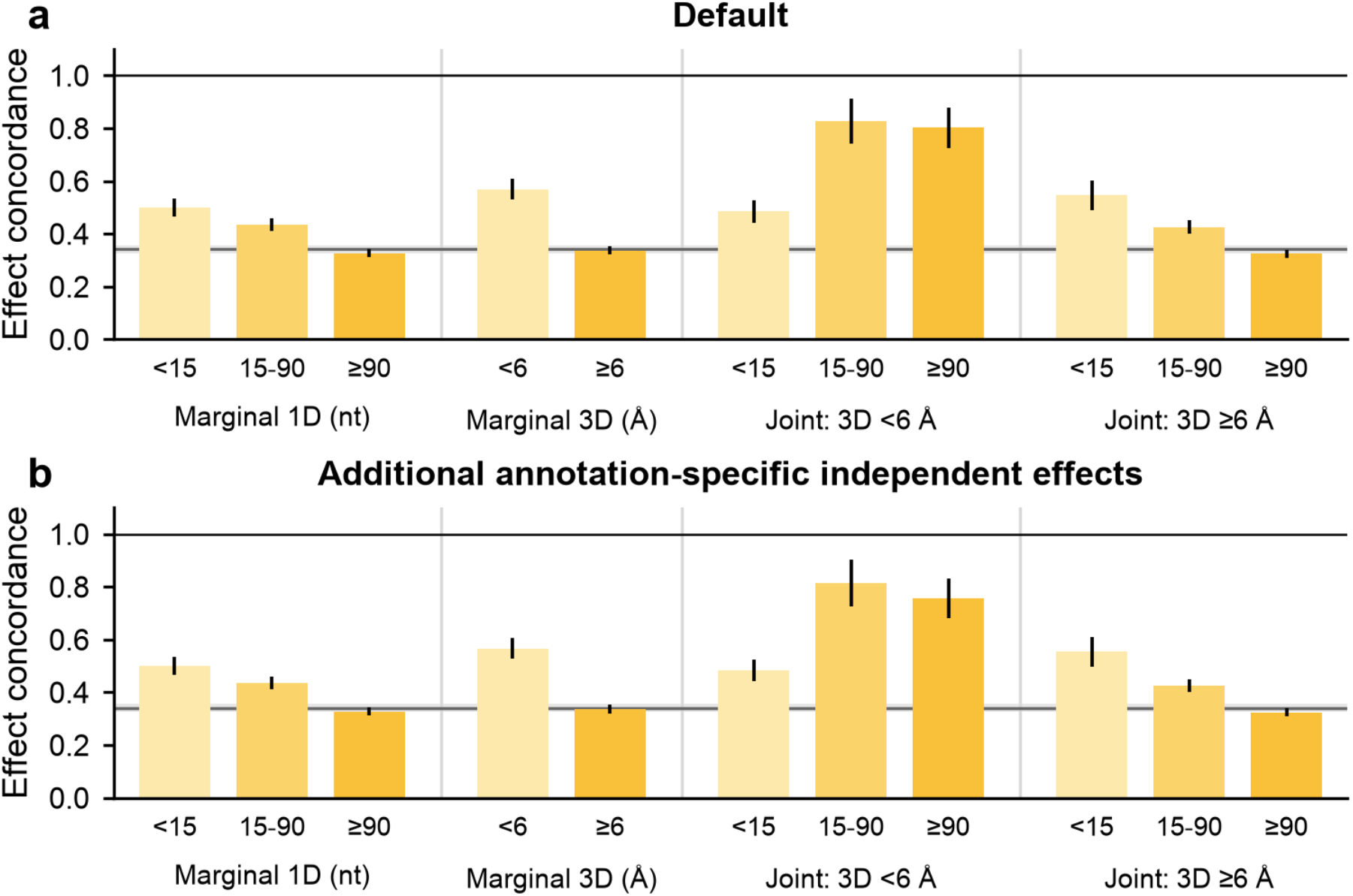
Effect concordance across 1D and 3D distances. Missense effect concordance (*ρ*) stratified by coding-sequence distance (<15, 15-90 or ≥90 nucleotides) or distance in AlphaFold-predicted structures (<6 or ≥6 Å), and their joint categories, **(a)** under the **Figure 6** model and **(b)** with an additional independent-effect kernel for variants in each pair set. Bars show pooled estimates across UK Biobank and All of Us traits. The gray line and band show the all-pairs concordance estimate ± standard error. Error bars show standard errors.

**Supplementary Figure 19.**
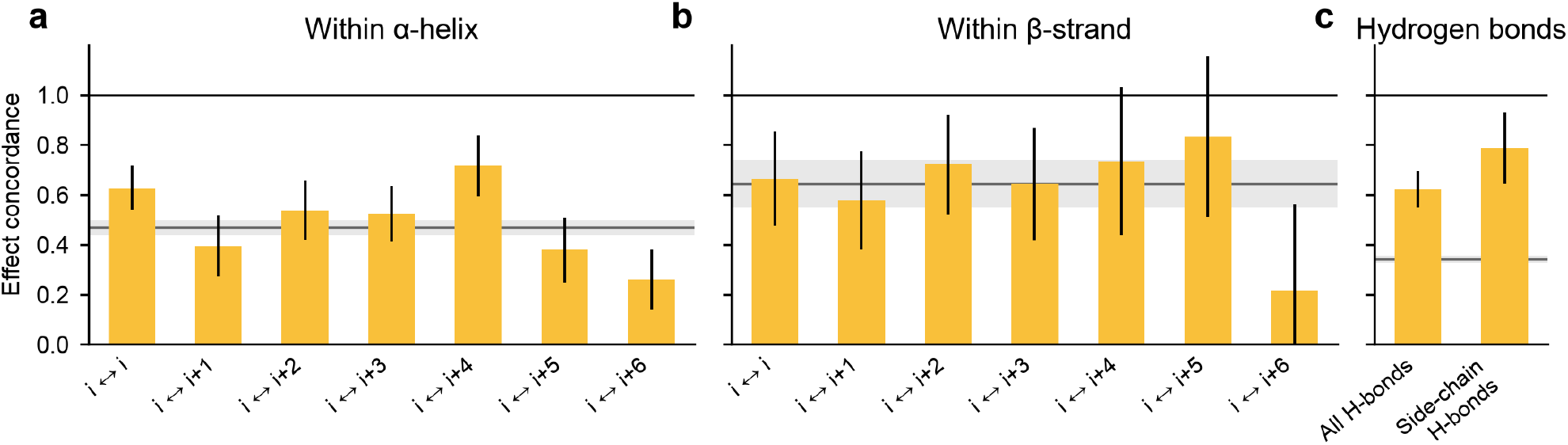
Effect concordance within secondary structures and between hydrogen-bonded residues. **(a,b)** Missense effect concordance by residue offset (0-6) within the same α-helix **(a)** or β-strand **(b)**; offset 0 denotes distinct variants at the same residue. Gray lines and bands show concordance ± standard error across all pairs within the element **(a**,**b)** or all within-gene missense pairs **(c**). **(c)** Concordance for all hydrogen-bonded residue pairs and pairs with at least one side-chain participant. Estimates are meta-analyzed across UK Biobank and All of Us traits. Error bars show standard errors.

**Supplementary Figure 20.**
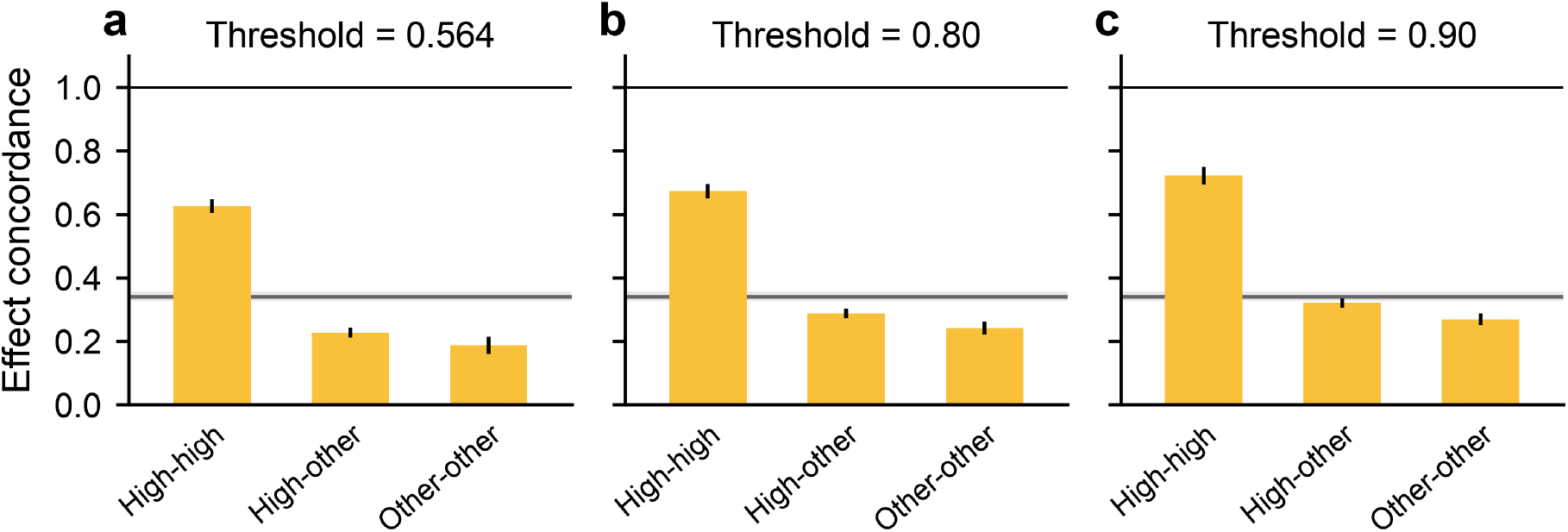
Effect concordance across AlphaMissense thresholds. Effect concordance among within-gene missense variant pairs at AlphaMissense thresholds of 0.564 **(a)**, 0.80 **(b)** and 0.90 **(c)**. Bars show high-high, high-other and other-other pairs, each fitted as a separate position kernel and estimated with an additional independent component for participating variants. Gray lines and bands show all-pairs concordance ± standard error. ‘High’ denotes scores above the threshold; ‘other’ denotes scores below the threshold. Error bars show standard errors.

**Supplementary Figure 21.**
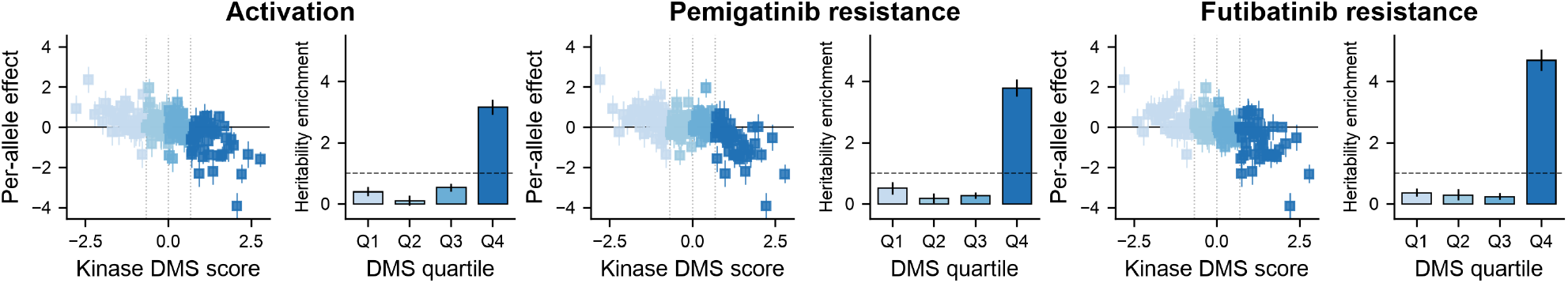
*FGFR3-*height allelic series across DMS experimental conditions. FGFR3 height effects and heritability enrichment using kinase-domain DMS scores measured under activation, pemigatinib-resistance and futibatinib-resistance conditions. The resulting patterns were consistent across experimental conditions. Within each cell, the left panel shows per-allele height effects against normalized kinase DMS scores, with blue shades denoting assay-specific score quartiles. The right panel shows heritability enrichment for the same quartiles, from the lowest (Q1) to highest (Q4) DMS scores. Error bars show standard errors.

**Supplementary Figure 22.**
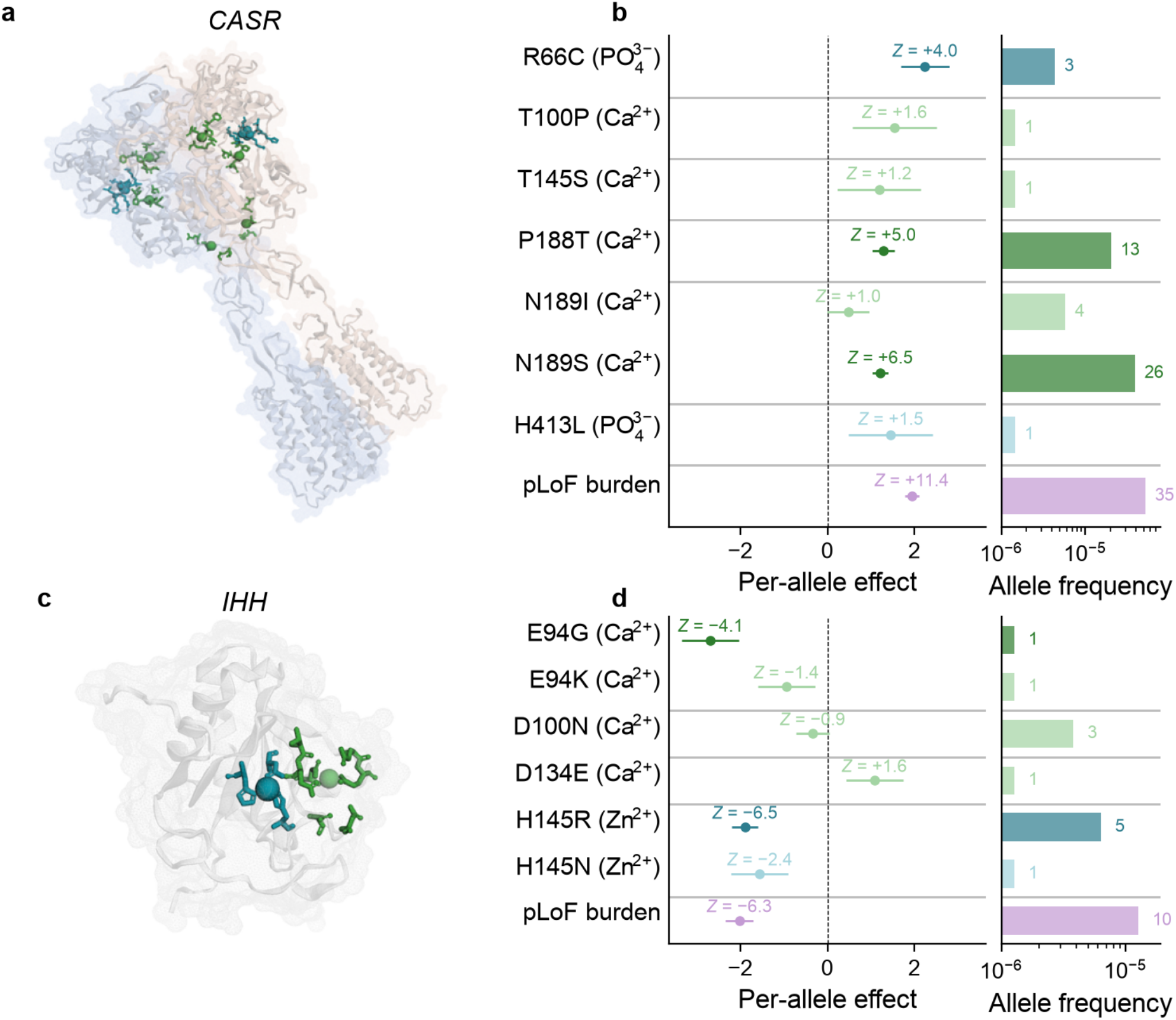
Allelic series in *CASR*-calcium and *IHH*-height associations. *CASR*-calcium: structural locations **(a)** and per-allele effects **(b)** of seven missense variants at six residues involved in calcium or phosphate binding. *IHH*-height: structural locations **(c)** and per-allele effects **(d)** of four variants at calcium-binding residues and two variants at the zinc-binding residue. Per-allele effects are shown with Z scores, allele frequencies and allele counts. Darker colors denote *P* < 0.001, and purple denotes the aggregate pLoF burden. Error bars show standard errors.

## Supplementary Tables

**Supplementary Table 1. Simulation for heritability estimation**. Numerical results for heritability estimation simulations (**Figure 1a**). Simulated estimands and mean heritability estimates with standard errors are reported across 100 replicates under three simulation settings.

**Supplementary Table 2. Simulation for heritability enrichment estimation**. Numerical results from the heritability-enrichment simulations (**Figure 1b**). Simulated estimands and enrichment estimates with standard errors are reported across 100 replicates under three simulation settings.

**Supplementary Table 3. Simulation in All of Us with no rare coding heritability**. Heritability estimates from All of Us simulations with common-variant effects but no rare coding variant effects. Means and standard errors of the mean across five replicates are reported for BHR-like, KASM Baseline, KASM Baseline x Exon and KASM Baseline + Indep.

**Supplementary Table 4. Exome-wide association statistics in UK Biobank and All of Us**. Phenotype definitions and sample sizes for 73 cohort-trait pairs, including 16 UK Biobank phenotypes in EUR participants and 19 All of Us phenotypes in each of the EUR-, AFR- and AMR-like genetic-similarity groups. Sample sizes are drawn from 394,841 exome-sequenced UK Biobank participants in Genebass^38^ and 392,030 All of Us participants in the All by All v8 release^39^.

**Supplementary Table 5. Correlations among missense scores and structural and functional annotations**. Pairwise Spearman correlations among missense scores and structural and functional annotations across missense variants.

**Supplementary Table 6. Functional and structural annotations**. Labels, sources, transformations and definitions for missense scores, structural and functional missense annotations, and synonymous scores.

**Supplementary Table 7. Rare coding heritability estimates across KASM models**. Random-effects, fixed-effects and mixed-effects heritability estimates with standard errors (**Figure 2b**) are reported for synonymous, pLoF and missense variants across the Naïve, BHR-like, KASM Baseline, KASM Baseline x Exon and KASM Baseline + Indep. models, for meta-analyzed and cohort-specific continuous traits and four UK Biobank binary traits on the observed scale.

**Supplementary Table 8. Rare coding heritability enrichment across gene-constraint quintiles**. Heritability enrichment estimates and standard errors (**Figure 2c**) across gnomAD pLoF observed/expected quintiles for pLoF, all missense and AlphaMissense-prioritized missense variants.

**Supplementary Table 9. Benchmark of missense deleteriousness predictors**. Within-gene top-decile heritability enrichment for individual missense scores and the KASM composite score for subthreshold and significant genes. Predicted mutation rate (Roulette-MR^102^) is included as a negative control; its top decile showed no enrichment. We report heterozygosity-normalized and count-normalized enrichment and heritability proportions with standard errors, and heterozygosity proportions.

**Supplementary Table 10. Deep mutational scanning benchmark results**. Quartile-specific deep mutational scanning and computational-predictor results for *FGFR3*-height, *GCK*-HbA1c, *LDLR*-LDL cholesterol and *MC4R*-BMI. We report heritability and heterozygosity proportions and enrichment with standard errors.

**Supplementary Table 11. Structural and functional enrichment of missense heritability**. Estimates for structural and functional protein annotations. We report heritability and heterozygosity proportions and enrichment with standard errors for subthreshold and significant genes.

**Supplementary Table 12. Context-dependent enrichment of missense heritability**. Numerical results among significant gene-trait associations across UK Biobank and All of Us. We report enrichment and standard errors for missense variants within and outside each structural or functional context, separately for variants with and without each substitution class, and two-sided P values for the difference.

**Supplementary Table 13. Effect concordance of rare coding variant pairs**. We report effect concordance with standard errors for missense and pLoF variant pairs across marginal and joint one- and three-dimensional distance ranges.

**Supplementary Table 14. Per-gene functional enrichment**. Per-gene enrichment results for significant gene-trait associations across UK Biobank and All of Us. We report heritability enrichment with standard errors, heterozygosity proportions, and one-sided P values for enrichment greater than one.

**Supplementary Table 15. *PIEZO1*-HbA1c allelic series**. Burden associations with HbA1c for *PIEZO1* interchain hydrogen-bond, all missense and pLoF variants, and heritability enrichment results.

**Supplementary Table 16. *FGFR3*-height allelic series**. Height heritability enrichment for *FGFR3* transmembrane-domain variants, all DMS-scored kinase-domain variants and each DMS quartile, with heritability and heterozygosity proportions, enrichment with standard errors and one-sided *P* values for enrichment greater than one.

